# Listening to Bluetooth Beacons for Epidemic Risk Mitigation

**DOI:** 10.1101/2021.01.21.21250209

**Authors:** Gilles Barthe, Roberta De Viti, Peter Druschel, Deepak Garg, Manuel Gomez-Rodriguez, Pierfrancesco Ingo, Heiner Kremer, Matthew Lentz, Lars Lorch, Aastha Mehta, Bernhard Schölkopf

## Abstract

During the ongoing COVID-19 pandemic, there have been burgeoning efforts to develop and deploy digital contact tracing systems to expedite contact tracing and risk notification. Unfortunately, the success of these systems has been limited, partly owing to poor interoperability with manual contact tracing, low adoption rates, and a societally sensitive trade-off between utility and privacy. In this work, we introduce a new privacy-preserving and inclusive system for epidemic risk assessment and notification that aims to address the above limitations. Rather than capturing pairwise encounters between user devices as done by existing systems, our system captures encounters between user devices and beacons placed in strategic locations where infection clusters may originate. Epidemiological simulations using an agent-based model demonstrate several beneficial properties of our system. By achieving bidirectional interoperability with manual contact tracing, our system may help reduce the effective reproduction number already at adoption levels of 10%. The use of location and environmental information provided by beacons allows our system to achieve significantly higher sensitivity and specificity than existing systems and thus may improve the efficacy of contact tracing under limited isolation and testing resources. Moreover, to achieve high utility, it is sufficient to deploy beacons in a small fraction of strategic locations. Finally, our simulations also show that existing systems could inherit these beneficial properties if they integrated the beacons used by our system.

## Introduction

Containing infectious diseases such as the ongoing COVID-19 pandemic requires effective testing, contact tracing, and isolation of infected individuals (TTI) [1–4]. Among these, contact tracing is an important tool that can help direct limited test resources to those most likely to be infected, by identifying infected individuals and their close contacts during the infectious period. Furthermore, contact tracing can provide insight into the circumstances of contagion, which in turn informs the implementation of public health policies and interventions to help contain disease spread. For instance, outbreaks of COVID-19 in meat packing plants in Germany [5] provided insights into conditions that can potentially breed infection hotspots, leading to regulatory action [6].

To expedite contact tracing, a number of digital contact tracing systems have been proposed [7–13]. In most of these systems, individuals install a smartphone application that records instances of physical proximity with devices of other individuals via close-range Bluetooth exchanges, referred to as encounters. Once an individual is tested positive, their recent encounter history is used to notify other at-risk individuals who have come into contact with the infectious person during her period of contagiousness.

Despite strong efforts to deploy these smartphone-based, pairwise encounter-based contact tracing systems, henceforth referred to as SPECTS, there is only limited evidence of their effectiveness [14–16]. This may be due to poor interoperability with manual contact tracing [14] and low adoption rates (*<* 25%) in countries that do not mandate their use [17], despite advertising campaigns to promote their adoption [18, 19]. Moreover, a societally sensitive trade-off between utility and privacy has precluded the use of location and associated environmental features (e.g., indoor vs. outdoor). This is a major disadvantage, since such measurements could help us predict individual risk of infection more accurately [20]. An in-depth overview of different SPECTS, including their adoption levels across countries, is provided in Section 4 of the *Supplementary Information*.

In the present work, we describe PanCast, a privacy-preserving, secure and inclusive system for epidemic risk assessment and notification that addresses the above limitations. In PanCast, Bluetooth beacons placed in strategic locations continuously broadcast ephemeral IDs. A subset of these beacons, called network beacons, also broadcast risk information associated with times when individuals who tested positive were near specific beacons. Individuals carry devices that listen to these beacons passively (i.e., without transmitting anything), store the beacons’ ephemeral IDs, and later compare the stored IDs against risk information broadcast by network beacons or received directly via the Internet. The user devices in PanCast can be inexpensive, zero-maintenance, small electronic dongles in the form of cards or key fobs, as well as more sophisticated devices such as smartphones.

When an individual tests positive for the infectious disease, they may be legally required to or choose to disclose (a selected subset of) the list of ephemeral IDs stored in their user device. The individual must explicitly authorize the transmission of data from their device via a trusted terminal. The trusted terminal can be part of a kiosk installed in a test center, clinic, or doctor’s office, or a personal device (e.g., a smartphone or a computer) owned by the user or a care provider. The information about which locations were contaminated at which times (which depends on users’ visits as well as on location features) then gets included in the broadcast risk messages. A user learns their risk of contagion due to their presence at sites visited by diagnosed individuals, but does not learn anything about locations visited by other individuals, unless these individuals decide to voluntarily disclose their locations.

Figure 1 shows an overview of PanCast’s architecture; cf. *Materials and Methods* and the *Supplementary Information* for more details on PanCast’s design, data collection and processing, risk dissemination, risk score calculation, and privacy and security properties. It is important to note that an alternative implementation of the system could use displayed QR codes [21, 22] instead of some or all of the non-network beacons. However, scanning a QR code requires explicit user action and unless QR codes are displayed using monitors and updated every few minutes, they end up being used for extended periods, which results in weaker security (see Section 4.2 of the *Supplementary Information*). Finally, PanCast’s beacons can broadcast ephemeral ids compatible with the Google/Apple Exposure Notification (GAEN) protocol used by most SPECTS [23], allowing the beacons to seamlessly interoperate with deployed apps.

**Figure 1:**
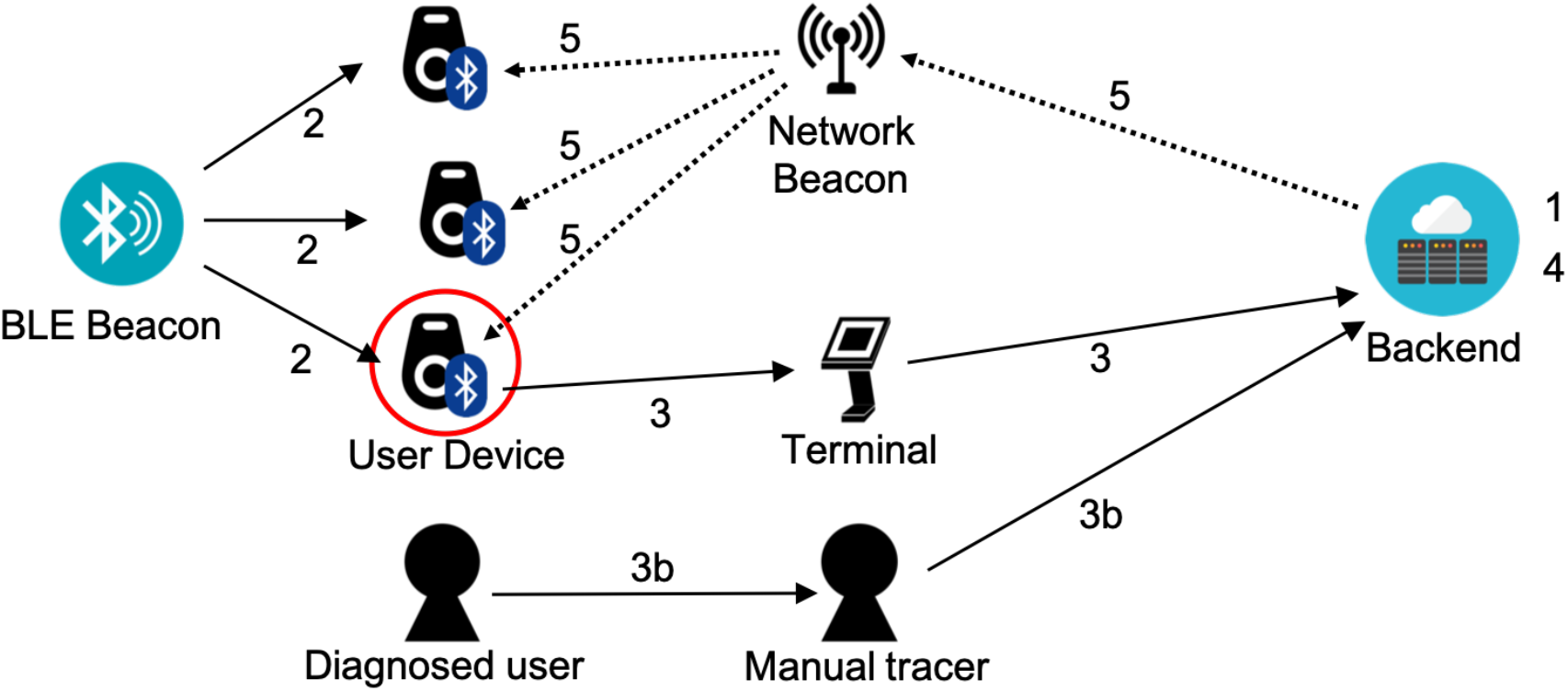
PanCast’s architecture. **1**. Beacons and user devices are registered with the backend. **2**. User devices record encounters with BLE beacons. **3**. Diagnosed users or healthy volunteers may upload their history of encountered beacons to the backend via a terminal. **3b**. Optionally, health workers can manually feed inputs from users into the backend system. **4**. The backend updates the risk database with uploaded encounters. **5**. Risk information is periodically broadcast from the backend to network beacons, which broadcast the information to nearby user devices.

By design, PanCast facilitates participation of technology-challenged, economically disadvantaged, or physically challenged individuals who cannot or do not wish to use smartphones. Moreover, it ensures data minimization in accordance with existing regulations for (manual) contact tracing—a healthy individual can use the system in a purely passive “radio” mode, individuals who test positive control which information they disclose (much like in a manual contact tracing interview), and this disclosed information is accessible only to individuals at risk and in a privacy-preserving way. Due to this passive mode, PanCast can achieve a similar level of privacy as existing digital contact tracing systems in spite of using location and environmental information provided by the beacons to enhance risk score estimation and identify infection clusters [24– 27]. Moreover, PanCast makes use of this information to achieve bidirectional interoperability with manual contact tracing: If an individual who tested positive uses a device, they can use the information saved in their device to better recall visited locations during a contact tracing interview, thus allowing PanCast to support manual contact tracing. Conversely, if a diagnosed individual did not use PanCast, a human contact tracer can manually create an entry in the risk database for any locations the individual recalls visiting, which may have installed beacons. By virtue of these properties, PanCast partially mitigates the so-called *x*^2^-adoption problem of SPECTS [12]: If a proportion *x <* 1 of the population has adopted a SPECTS, encounters get noticed with probability *x*^2^ ≪ 1.

To evaluate the performance of our system, we perform epidemiological simulations using an agent-based model [28]. Our results suggest that, by achieving bidirectional interoperability with manual contact tracing, the system helps reduce the effective reproduction number even under relatively low adoption. Further, our results show that by utilizing environmental information to improve risk estimation and inform tracing decisions, PanCast can achieve significantly higher specificity for the same levels of sensitivity when compared to a generic version of SPECTS. This can reduce the number of unnecessarily quarantined individuals, and in the case of limited isolation and testing capacities, it can improve epidemic risk mitigation by more efficient resource allocation. Moreover, our results also show that, to achieve high utility, it is sufficient to deploy beacons in a small fraction of strategic locations. Finally, our epidemiological simulations suggest that the above mentioned properties can benefit deployed SPECTS if PanCast beacons broadcast GAEN- compatible information. More specifically, the location information provided by PanCast can augment the information collected by SPECTS, enabling more accurate methods to increase precision and specificity of their notifications and enabling them to interoperate with manual contact tracing.

## Results

We simulate the deployment of our system exemplarily in Tübingen, a town with 90,546 inhabitants in south- west Germany. Individuals move between their homes and 1,479 different points of interest (POIs). In the simulations, we place beacons at five types of sites and identify their real locations using OpenStreetMap [29]: (i) schools, universities and research institutes; (ii) restaurants, cafes and bars; (iii) bus stops; (iv) offices and shops; and (v) supermarkets and convenience stores. To model the exposure of individuals at these sites, we use a recently introduced agent-based and POI-based epidemiological model [28]. The model quantifies the influence that individual mobility patterns, environmental drivers, as well as TTI have on the rate of transmission of each infected individual.

To compare the efficacy of PanCast with generic smartphone-based, pairwise encounter-based contact tracing systems (SPECTS), we simulate a range of epidemiological scenarios in which either PanCast or a generic SPECTS is employed over a six-month period. All experiments build on the same basic setup: starting with a completely susceptible population, we simulate an influx of infected individuals by randomly infecting 5 individuals per 100,000 inhabitants per week during the whole simulation period. Since our goal is to evaluate and compare the reduction in the number of infected individuals and the reproduction number achieved by PanCast and SPECTS, for simplicity, we do not deploy any additional interventional measures. In practice, to achieve epidemic control (*R*_eff_ *<* 1), one may need to deploy such additional measures, as argued elsewhere [3, 30–32]. However, our findings may also apply to settings in which additional measures are in place.

*Materials and Methods* provide further details on the epidemiological model used for the simulations as well as the implementation of the various tracing measures. If not stated otherwise, the results presented below are averages of 100 random roll-outs of the simulation, and the error bars correspond to one standard deviation. Section 5.2.3 of the *Supplementary Information* provides further analyses exploring the possibility of interoperation between our system and SPECTS. In this context, we find that a generic SPECTS with an adoption level between 10% to 25% already benefits from integrating PanCast beacon information from only a small proportion of POIs.

### Interoperation with manual tracing can improve efficacy at low adoption levels

To explore the effect of PanCast’s interaction with manual contact tracing, we combine either PanCast or SPECTS with manual contact tracing in a scenario where transmission rates are independent of the site type, i.e., the infection probability of a susceptible person does not depend on the place where the contact with an infected individual happened. This allows us to explore the effect of interaction with manual tracing independent of PanCast’s advantage of utilizing environmental information. While SPECTS and manual contact tracing operate independently of each other, PanCast and manual contact tracing can benefit from each other by exchanging information (see *Materials and Methods* for details). For all systems, tracing is initiated whenever the inferred exposure risk of a person exceeds the threshold of a 15 minute contact with an infected individual. We assume sufficient testing and isolation capacities for all individuals selected for tracing by a digital system or via manual contact tracing.

Figure 2 visualizes different aspects of PanCast and SPECTS in the presence of manual tracing. Figure 2a) shows the reduction of the number of infections achieved by the tracing systems with respect to the baseline scenario in which only manual contact tracing is employed. By mitigating the *x*^2^-adoption problem via interoperation with manual tracing, we observe that PanCast generally scales more favourably with decreasing adoption levels. The results suggest that in the ideal scenario where beacons are placed at all sites, an adoption level of roughly 20% with PanCast would be sufficient to reduce the number of infections by 10% compared to the baseline, whereas the same result would require around 50% adoption with SPECTS. We next investigate how the performance of PanCast decreases when beacons are placed strategically only at a subset of the sites. To this end, we place the beacons at a proportion of sites with the highest integrated visit time, defined as the sum of individual visit durations at a given site over a period of one month. Figure 2a) shows that for each proportion of sites with beacons, there is an adoption level up to which PanCast turns out to be more effective than SPECTS. With beacons at 25% of the sites (370 beacons in Tübingen) PanCast shows a clear advantage over SPECTS up to adoption levels of more than 50%. We observe that in order to surpass the efficacy of the SPECTS currently employed in Germany with an adoption level of 29% [33], deploying PanCast with beacons placed strategically at roughly 10% (148 beacons in Tübingen) of the sites would be sufficient. Finally, equipping only roughly 5% of the sites with beacons would be sufficient to match the performance of SPECTS for adoption levels of up to 25%, which is well above the adoption levels currently observed in many countries [34]. Figure 2b) shows the number of infected individuals over time at 25% and 50% adoption of digital tracing, respectively. We observe that the cumulative and peak numbers of infected vary across different strategies. Note that in scenarios with stronger outbreaks, herd immunity, defined as a sufficient number of immune individuals such that the effective reproduction number is below 1, will be reached at an earlier time, which explains the relatively larger numbers of infected in the later stage of the simulation period in scenarios with more efficient contact tracing. The left panel in Figure 2c) shows the effective reproduction number *R*_eff_ during the first weeks of the simulation averaged up until the point at which 10% of the population are not susceptible anymore, i.e., before the first signs of herd immunity set in. We observe that in our simulation with only manual contact tracing, *R*_eff_ is approximately 2.0 and decreases with increasing adoption of the digital tracing technologies to a minimum of approximately 1.3.

**Figure 2:**
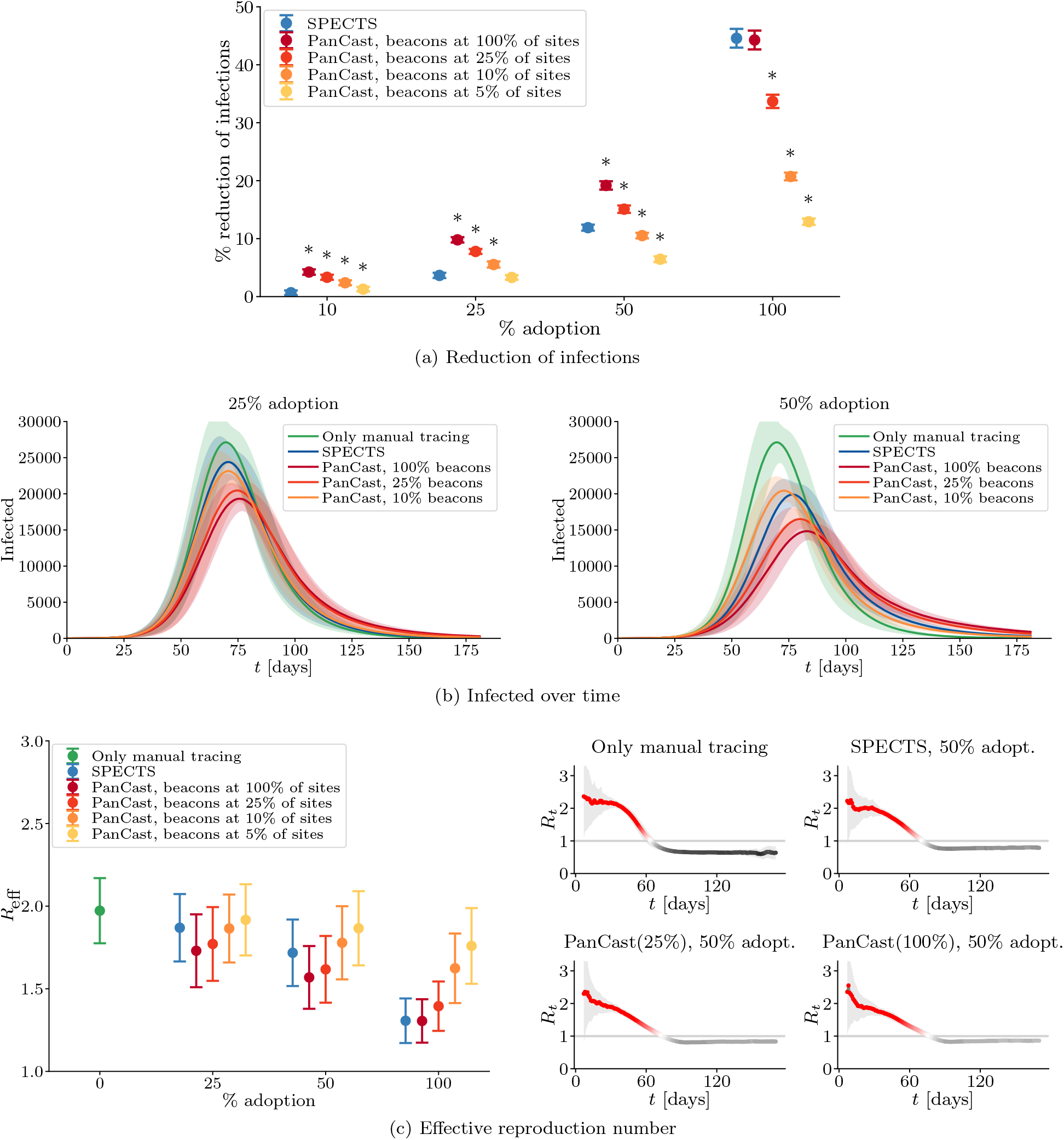
Interoperation with manual contact tracing. All experiments operate manual contact tracing and digital tracing in parallel. In contrast to SPECTS which do not interact with manual contact tracing, PanCast and manual contact tracing can benefit from each other by sharing information and can thereby improve the efficacy of the contact tracing efforts especially at low levels of adoption. Figure a) shows the reduction of infections and Figure b) the number of infected individuals over time. In Figure a), the sign ^*^ indicates statistically significant differences (two-sample t-test; p-value *<* 0.05) between PanCast and SPECTS. The left panel in Figure c) shows the effective reproduction number averaged over the period before more than 10% of the population are not susceptible anymore and the effects of herd immunity become relevant. The right panel shows the reproduction number of different scenarios over time. Lines and points represent averages of 100 random roll-outs of the simulation, error bars correspond to plus and minus one standard deviation.

The effective reproduction numbers are consistent with Figure 2a): PanCast leads to lower values of *R*_eff_ at low adoption, and for each value of the proportion of sites with beacons there is a level of adoption up to which PanCast seems to be favorable. The effective reproduction number over time is shown for four different scenarios in the right panel of Figure 2c).

Section 5.2.1 of the *Supplementary Information* provides further analysis of the interaction between digital and manual contact tracing. These include results for PanCast and SPECTS in the absence of manual tracing, the influence of delays in the manual tracing process, and a sensitivity analysis for the manual contact tracing parameters.

### Utilization of environmental information improves tracing accuracy

Environmental factors such as indoor vs. outdoor, room size, ventilation, and air quality, have been shown to lead to vastly different transmission rates [35]. To evaluate the effect of using such environmental information to improve contact tracing decisions, we simulate scenarios in which different site types have different transmission rates. As an example, we assume contacts with infected individuals at social sites, i.e., restaurants, bars and cafes to be ten times more likely, and contacts at bus stops to be ten times less likely to lead to infection than the default. Further, we normalize all (scaled) transmission rates by the same empirical factor such that the overall course of the epidemic remains invariant under these scalings. Unlike SPECTS, PanCast has access to environmental information and can therefore use the known variable transmission rates to estimate infection probabilities in our simulations. The SPECTS simulation uses an average transmission rate to calculate infection probabilities and make tracing decisions.

Figure 3 visualizes the results of several different experiments in this general setting. Figure 3a) shows the effect of utilizing environmental information for contact tracing on the course of the epidemic relative to the no contact tracing baseline. We assume that at any given time we allow a maximum of 10% of the population to be quarantined based on tracing decisions, in addition to positively tested individuals and their household members. Under this fixed budget, PanCast and SPECTS have to allocate tests and quarantine measures based on their respective assessment of infection probabilities. To isolate the effect of environmental information from PanCast’s interaction with manual contact tracing, we first employ both tracing systems in the absence of manual contact tracing (left panel). We observe that while SPECTS seems to provide a slightly stronger reduction in the number of infected at 10% adoption, PanCast with beacons at only 10% of sites achieves a larger reduction for higher adoption values. Furthermore, we see that 5% of sites with beacons seems to be sufficient to exceed SPECTS performance at 50% adoption. For adoption levels above 25%, equipping only 10% of the sites with beacons does not seem to negatively affect PanCast’s performance compared to placing beacons at all sites. The right panel of Figure 3a) shows the results of the same experiments in the presence of manual contact tracing. We observe that by leveraging manual contact tracing information, both PanCast and SPECTS achieve higher levels of reduction of the number of infected compared to the results achieved without manual tracing, especially at low adoption levels. Moreover, the results suggest that PanCast’s advantage over SPECTS increases via its interaction with manual contact tracing, for proportions of sites with beacons at 10% and above. Section 5.2.2 of the *Supplementary Information* provides further analysis concerning the utilization of environmental information. We find that the relative advantage of PanCast is consistent across alternative isolation capacities.

**Figure 3:**
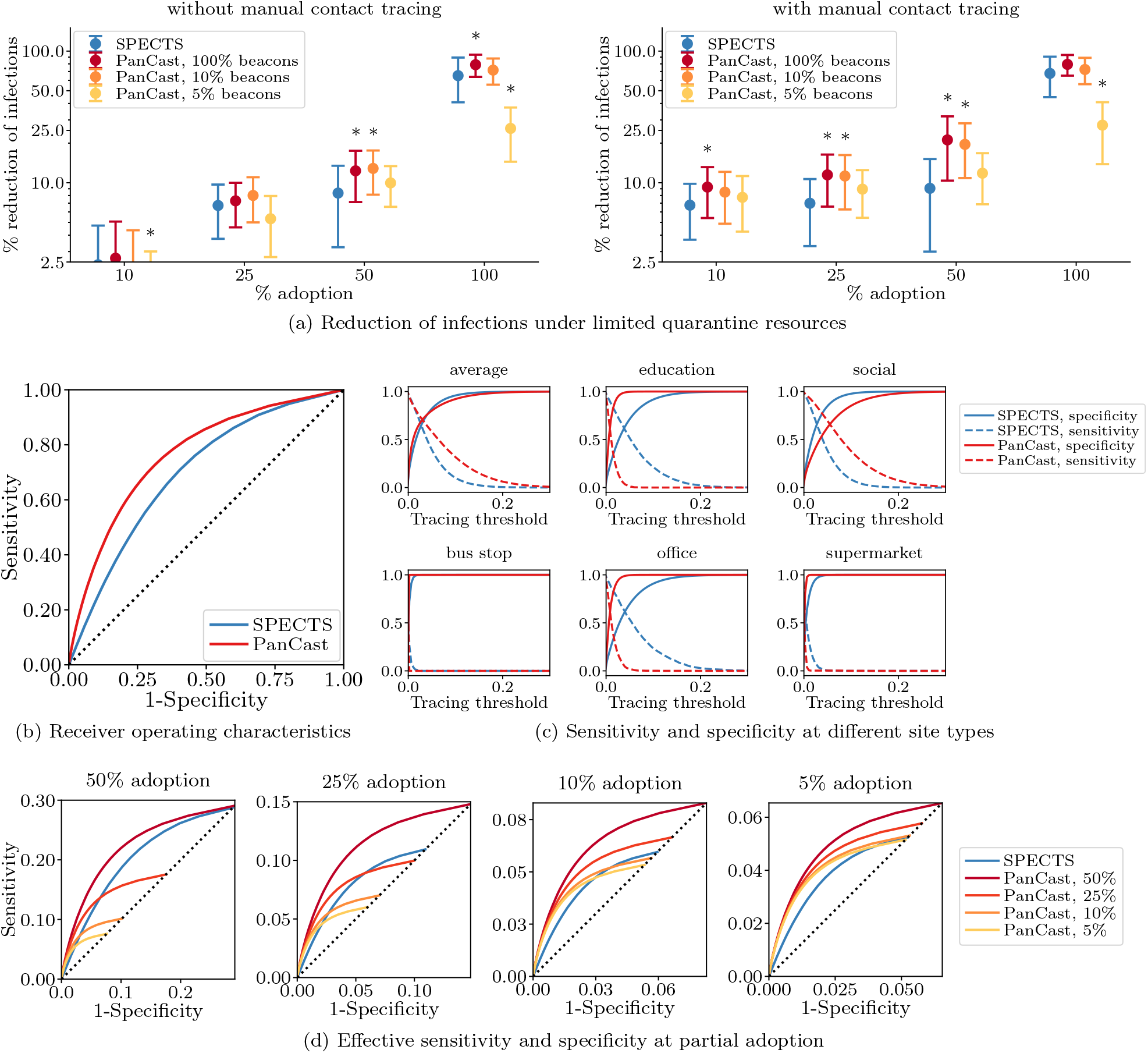
Leveraging site information to improve tracing decisions. We assume that PanCast has access to the site-dependent transmission rates, while SPECTS can only use an average value to inform tracing decisions. Figure a) shows the reduction of infections achieved by PanCast and SPECTS with and without manual tracing under the constraint that at any given time a maximum of 10% of the population can be quarantined due to tracing decisions (in addition to positively tested individuals and their household members). In both figures, points represent averages of 400 random roll-outs of the simulation, error bars correspond to plus and minus one standard deviation, and the sign ^*^ indicates a statistically significant difference (two-sample t-test; p-value *<* 0.05) between PanCast and SPECTS. Figure b) shows ROC curves defined as true positive rate (sensitivity) against false positive rate (1 – specificity). Figure c) stratifies sensitivity and specificity by site type. Figure d) shows *effective* sensitivity and specificity (see text) for different adoption levels, incorporating both interaction with manual contact tracing and utilization of environmental information. PanCast outperforms SPECTS if adoption is low or the percentage of sites with beacons is high.

Figure 3b) compares PanCast’s and SPECTS receiver operating characteristic (ROC) curves computed by varying the tracing threshold, i.e., the infection probability above which contacts are selected for tracing, from 0 to 1. We observe that for every fixed sensitivity (true positive rate) PanCast achieves a larger specificity (i.e., smaller false positive rate), implying that PanCast can detect the same proportion of infected individuals as SPECTS while imposing a smaller burden on the population in terms of a smaller number of unnecessarily quarantined individuals.

Figure 3c) shows the sensitivity and specificity of the two tracing systems against the tracing threshold for different site types. We see that on average PanCast provides larger sensitivity and approximately equal specificity compared to SPECTS for every value of the threshold. Further, we observe that SPECTS overestimate the infection probability at education, office and supermarket sites while underestimating it at social sites, which explains the larger false positive rate for a given true positive rate as compared to PanCast. Notice that, resulting from our rescaling of the site-dependent transmission rates, bus stops and to a lesser degree supermarkets become (almost) irrelevant for the epidemiological development, as transmission rates are low and contact times between individuals typically short. As the number of infections at these sites tends to zero, specificity and sensitivity are not well defined anymore.

Figure 3d) visualizes the efficacy of the contact tracing systems in a different setting, where system adoption is taken into account. More specifically, while sensitivity and specificity are initially only defined over the group of individuals who participated in the study, i.e., only contacts for whom the tracing system was capable of making an active decision for or against tracing, we define the *effective*, also called *clinical* [36], sensitivity and specificity as taking into account all individuals in the simulation. For all individuals that could not be traced due to either the infector or the potentially infected individual not participating in contact tracing, we assume a negative tracing decision. We study the *effective* quantities in the presence of manual contact tracing, thereby combining the aspects of both interoperation with manual tracing and utilization of environmental information. We observe that PanCast generally outperforms SPECTS if adoption is low or the percentage of sites with beacons is high. If neither is the case, then PanCast still has higher sensitivity up to certain values of the false positive rate (i.e., 1 specificity), above which the situation is reversed. Finally, in real-world application, the relative advantage of PanCast may further increase as more fine-grained and accurate information characterizing the infection risks of sites becomes available, allowing the use of data-driven infection risk estimates [37]. While such knowledge is presently scarce, a system such as PanCast would allow us to gather it.

### Strategic placement of beacons allows for high utility at low cost

In the previous experiments, we have placed beacons strategically at sites where exposures are most likely to happen. Figure 4 compares this to a random placement strategy within the scenario of site-independent transmission rates (i.e., the scenario of Figure 2). The results in Figure 4a) show that a random placement would require equipping a large proportion of roughly 50% of the sites with beacons to achieve at least 10% reduction in the number of infections for low and intermediate adoption levels. This disadvantage is mitigated by placing beacons at the sites with the highest integrated visit times. In the latter case, we observe that even for proportions of sites with beacons below 25% (i.e., 370 beacons in our Tübingen example) PanCast is capable of reducing infections by at least 10% for low and intermediate adoption levels. Figures 4b) and 4c) show the distribution of beacons for the two strategies. We observe that the random strategy places many beacons at offices and workplaces (red) while the other strategy places beacons predominantly at social (yellow) and education sites (blue). Note that these results depend on the mobility model, and with another model, different sites or site types could be prioritized. Finally, note that these estimates are conservative as we ranked the sites only according to integrated visit time and did not account for differences in site-specific transmission rates. Any features known to affect infection risk (e.g., indoor vs. outdoor) could be used to further improve beacon placement.

**Figure 4:**
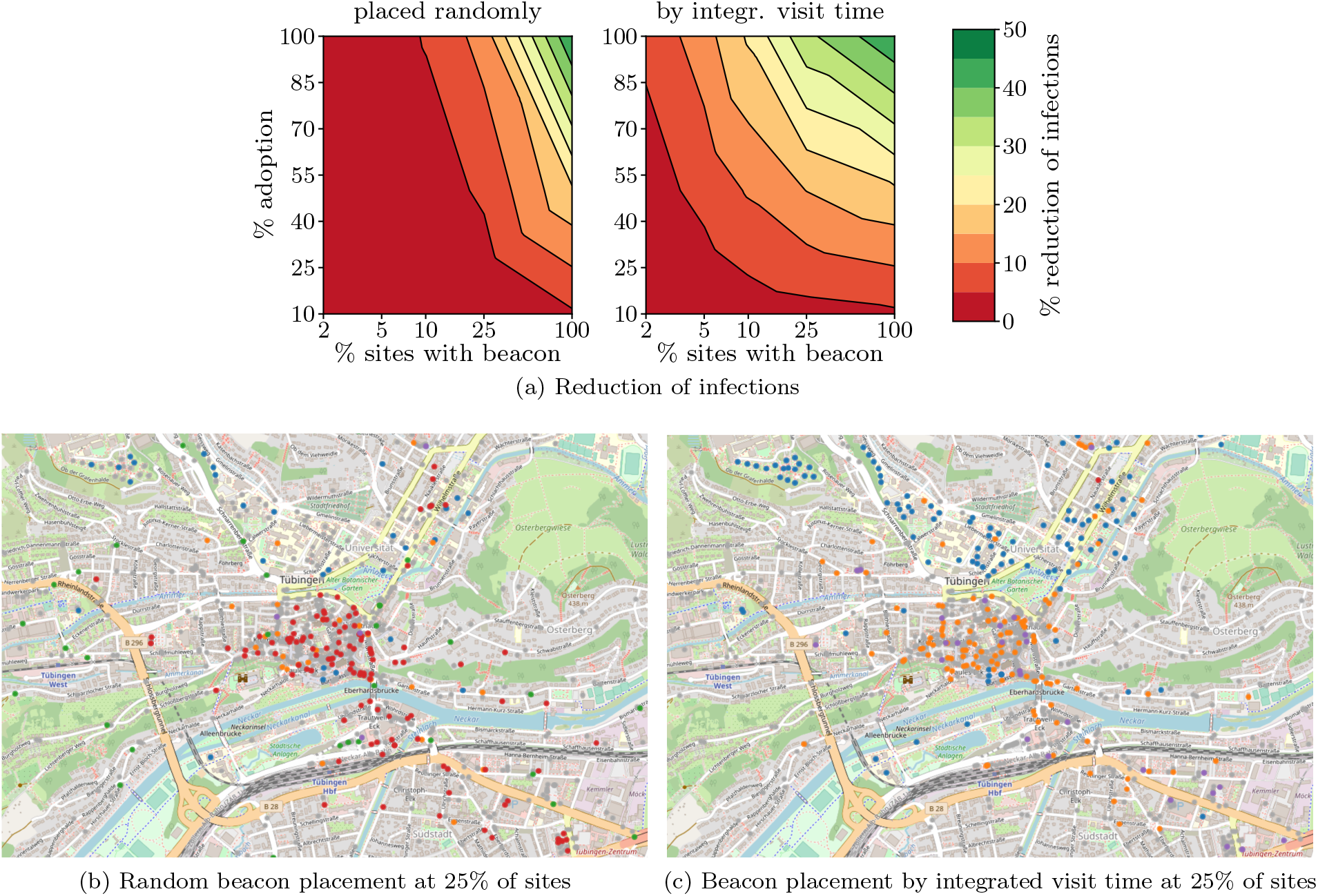
Beacon placement strategies. Figure a) shows the reduction of the number of infections under random and strategic allocation of beacons respectively over the proportion of beacons and the adoption level of PanCast averaged over 100 simulations. Figures b) and c) show the spatial distribution of beacons at 25% of the sites across our example city, Tübingen. Colored circles mark sites equipped with beacons and grey circles represent sites without beacons. The site types in our model are shops and workplaces (red), cafes, bars and restaurants (orange), schools and universities (blue), grocery stores (purple) and public transport stops (green).

## Discussion

While manual contact tracing has played an important role in epidemic mitigation strategies [14, 38], digital tracing solutions have not been able to prove their efficacy in countries that do not mandate their use, partly due to low adoption levels [15, 16, 18]. As shown by the simulations, bidirectional integration of digital and manual tracing has the potential to yield benefits for epidemic mitigation at relatively low adoption levels. Further, our results suggest that the utilization of environmental information for risk assessment can significantly improve the efficacy of contact tracing decisions and thereby contribute to effective mitigation. While existing SPECTS can neither effectively interact with manual tracing nor take into account environmental information, this limitation could be relaxed if PanCast’s beacons implemented SPECTS protocols, and the information provided by SPECTS is combined with the information provided by the beacons. This applies both for centralized SPECTS such as PEPP-NTK [7] and decentralized ones such as DP3T [11]. By doing so, PanCast could add relevant location-dependent information to SPECTS, enabling them to interoperate with manual contact tracing and increasing the accuracy of their notifications. In addition, any information obtained during manual contact tracing of individuals who tested positive could be used to populate the PanCast risk database. One could further implement proactive contact tracing [37, 39] on PanCast, i.e., not only trace the contacts of positively tested individuals but also the contacts of their contacts.

In addition to aspects concerning the interoperability with other systems, effective digital tracing should be able to both leverage and help acquire circumstantial information that influences exposure. It has recently been shown that environmental factors such as indoor vs. outdoor spaces, room size, and ventilation strongly influence individual risk of contagion [20]. By placing beacons in known locations, PanCast can account for these environmental factors to achieve more accurate estimates of individual risks of infection, as shown in our epidemiological simulations. If required, one may estimate some factors such as distance more precisely using triangulation among multiple beacons per location [40]. Moreover, while SPECTS are limited to contemporaneous encounters where individuals occupy a space at the same time, PanCast can capture non-contemporaneous transmissions, which may occur shortly after an infectious individual has left a small and poorly ventilated room. While our simulations do not explicitly show this advantage due to a lack of reliable estimates for the true infection risk as a function of space-time distance, there are reasons to believe that non-contemporaneous infections contribute to the course of the pandemic [41]. A deployment of PanCast would allow for gathering the data needed to estimate these or other disease parameters, which could subsequently be used to evaluate the system and assist research. This also concerns the *overdispersion* of infections observed for COVID-19—many infected individuals do not infect anyone, while few *superspreaders* infect many [24–27]. Therefore, while a diagnosed patient is unlikely to be a superspreader themselves, they are likely to have been infected by a superspreader who has probably infected others as well. If digital contact tracing can identify infection events spatially, one may be able to trace entire clusters using such backward contact tracing [39, 42].

While PanCast exhibited favorable results in the simulations, the conclusions we draw have to be considered in the context of the epidemiological model employed, similarly as in previous work assessing the effectiveness of SPECTS [4, 39, 43]. In this context, it is worth noting that, in our simulations, we did not consider additional interventional measures. As a result, a large proportion of the population becomes infected and the epidemic is eventually ended by a combination of contact tracing and herd immunity. However, our findings suggest that, as PanCast can lead to a stronger reduction of the reproduction number, it may be possible to achieve epidemic control (*R*_eff_ *<* 1) under less stringent interventional measures than with SPECTS. Moreover, while we have used a suitable modeling framework, explicitly representing the sites at which transmissions occur, the simulations of both SPECTS and PanCast do not take into account practical considerations such as battery status or imperfect beacon coverage within a specific site. The simulations moreover assume that risk dissemination is instantaneous. This assumption holds for users carrying smartphones, which can frequently query and download new risk information via the Internet. For users carrying dongles, the assumption is reasonable if such individuals’ movement is mostly local, which is a good approximation especially while individuals’ movement is constrained by government interventions [44]. It would be unrealistic if individuals frequently traveled to distant sites, in which case network beacons around the world would have to broadcast comprehensive non-local risk information which takes more time. However, our detailed protocol for risk dissemination (Section 2 in the *Supplementary Information*) ensures that even if individuals travel to far away sites, the time it takes for dongles to receive risk information stays within acceptable limits.

Ultimately, digital tracing can only be effective if it can be easily deployed, is widely accessible and available at low cost, and does not suffer from delays. While PanCast can be implemented using smartphones as user devices, the use of dongles as user devices also allows individuals who, for financial or personal reasons, cannot or do not want to use smartphone apps to participate in and benefit from digital contact tracing. As these individuals might be overrepresented among the elderly and socio-economically disadvantaged, PanCast specifically allows for the inclusion of groups that may be particularly vulnerable to pandemics [45]. While a broad installation of PanCast would require the distribution of dongles to (at least) individuals without smartphones, as well as equipping a significant amount of sites with beacons, the system could also be effectively employed in an incremental and/or local fashion at low costs. Individual institutions such as schools or companies could employ the system on their premises to trace and control the epidemic spread among their members. Such local deployment would already provide valuable information on location-dependent transmission parameters. In addition, it would allow tracking these parameters over time as a virus may change its characteristics in response to the evolutionary pressure generated by containment measures. This has the potential to contribute to a more fine-grained, data-driven understanding of pandemics.

## Materials and methods

### System components

Figure 5 shows an overview of PanCast’s architecture. PanCast comprises two types of Bluetooth Low Energy beacons (BLE-only and BLE+network), user devices (dongles or smartphones), terminals, and a backend platform that relays risk notifications and aggregates data for epidemiological analysis. In the rest of this section, we mainly focus on dongles for user devices. All beacons and dongles are registered and authenticated with the backend, receive a secret key from the backend at the time of registration, and have a coarse-grained timer and a small amount of flash storage. When a user receives a dongle, they receive a list of one-time passwords (OTPs) that are also stored in the dongle. A diagnosed user can use these OTPs to authenticate to the dongle and to control the upload of data. Below, we elaborate on the components of the system and their respective functionalities.

**Figure 5:**
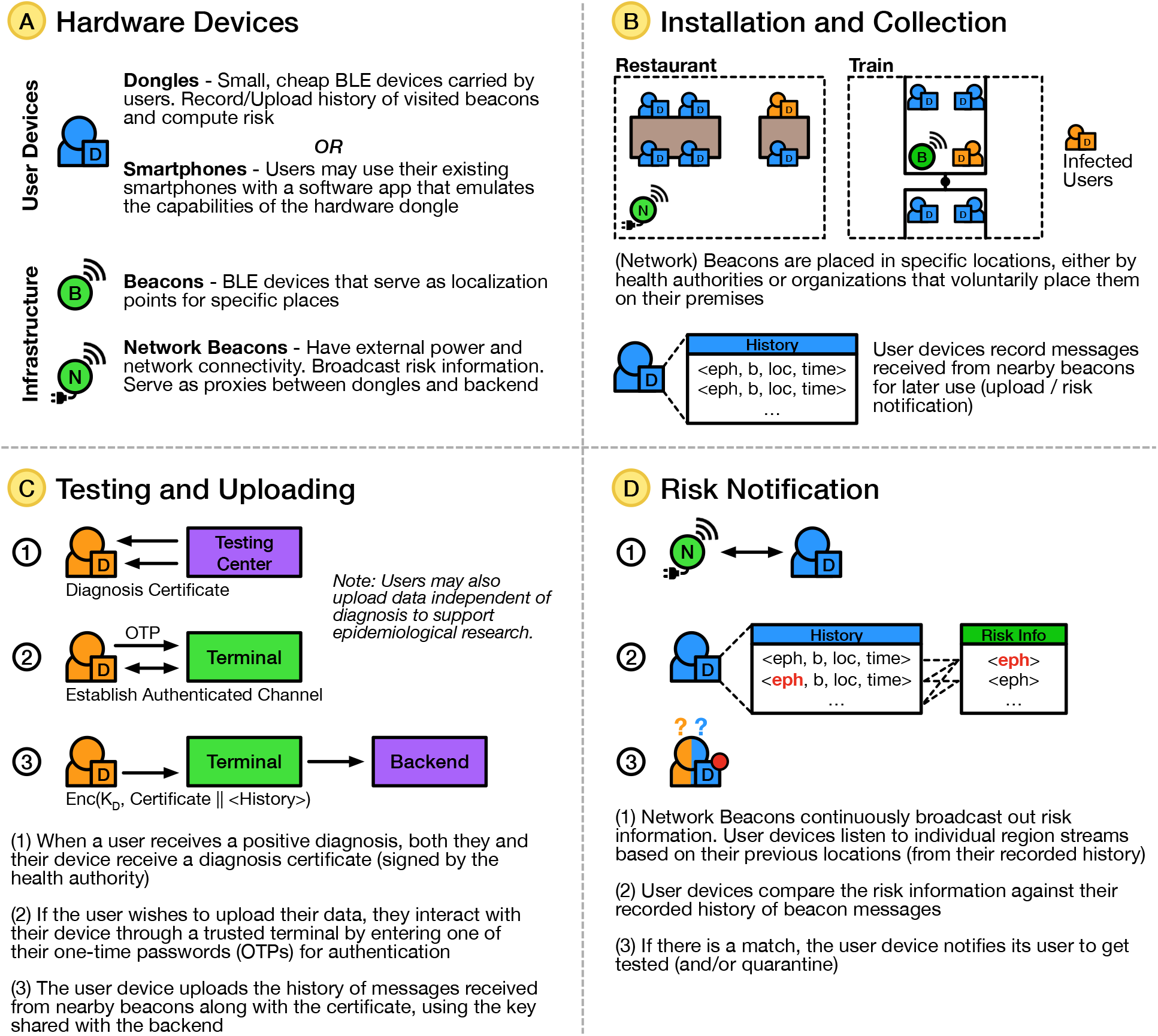
PanCast’s hardware devices, installation and collection, testing and uploading, and risk notification.

#### Beacons

PanCast employs two types of beacons: *BLE beacons* are commodity, battery-operated BLE-only beacons and require no network connection to the backend. *Network beacons* also use BLE, but additionally require mains power and a network connection to provide connectivity to backend servers. The beacons serve two purposes:

i. Every beacon provides a localization point in a specific place (e.g., an office, a bar, a public bus). A beacon periodically broadcasts an ephemeral id, its device id, and a location id. The device id and the location id are cryptographically signed by the backend, while the ephemeral id is generated by the beacon by hashing a beacon-specific secret key, its location id, and an epoch number derived from its local clock. Because the hash includes the epoch number, a fresh ephemeral id is broadcast in every epoch. This localizes every encounter between the beacon and a dongle to a specific epoch. The epoch length is set upfront to a small value, e.g., 15 minutes.
ii. Network beacons additionally broadcast global risk information received from the backend periodically, using a protocol that balances privacy and efficiency.

Beacons are installed in specific locations, for instance, under the guidance of health authorities or by organizations that choose to place them on their premises. Stationary beacons broadcast a GPS coordinate or a named identifier (e.g., city, zip code). Beacons may also be installed in mobile locations such as trains or buses; in this case they broadcast ids that identify their trajectories. All beacons are registered with the backend using their id and their location id comprising their stationary coordinate or trajectory, as well as information about their location that may be epidemiologically relevant (e.g., indoor, outdoor, ventilation, air quality, ambient noise level). This information can be used by the backend when computing infection risks or performing epidemiological analyses. Simple, battery-operated BLE beacons can account for the majority of beacons. These can be cheap and easy to install, because they do not require mains power or network connectivity. We expect them to be installed wherever infection transmission is likely to occur (e.g., in places where people congregate). A smaller number of network beacons provide nearby dongles with risk information. To reduce installation costs, we expect them to be installed where power and network connectivity is already available, e.g., next to WiFi base stations.

#### Dongles

Dongles are small, simple devices that users can attach to a keyring, or wear on the wrist or around the neck. They operate off a coin battery and have a minimal user interface in the form of a LED that indicates risk status and battery condition, and a button to control the LED notification. Dongles continuously listen for BLE transmissions from nearby beacons. They receive ephemeral ids from both types of beacons and store them along with the timestamps. When in proximity of a network beacon, they additionally receive risk information, compare that information with the device’s stored history of ephemeral ids received from beacons, and alert the user in case they were in spatio-temporal proximity of a diagnosed individual under circumstances that suggest a possible transmission. Dongles usually operate entirely passively. They only transmit information when a user chooses to anonymously reveal their information via a user terminal.

#### Backend service

The backend maintains several databases. First, it maintains a database of registered beacons, their locations/trajectories, and the secret key used by each beacon to generate the unique sequence of ephemeral ids it broadcasts. A second database contains registered users, their dongles, and the cryptographic keys required to authenticate each dongle. A third database, called the risk database, contains the uploaded encounter histories of individuals who recently tested positive. Finally, a fourth database, called the epidemiology database, contains the encounter histories of healthy users who chose to contribute their data for epidemic analytics. The encounter database is available to health authorities for analytics, e.g., to identify hotspots, superspreading events, and to estimate epidemiological parameters. The backend uses this database to transmit risk information to network beacons, which then broadcast it to nearby user dongles.

#### User terminals

User terminals are provided at locations that issue dongles as well as health care facilities that do testing. Terminals allow users to connect to their dongles over BLE to change their privacy settings, inspect what data is recorded on their dongles, upload data to the backend and, when allowed or required by law, decide what subset of the recorded information they wish to upload when they are tested positive. Users can also use personal computers or smartphones as terminals to perform these tasks in the comfort of their home or use the smartphone of a care provider who visits their home.

### System functionality

#### Data upload and risk dissemination

Diagnosed individuals may share their encounter data with health authorities to enable dissemination of risk information. Upon receiving a positive test result, users obtain a certificate with which they can verify their infectious state when uploading the information stored in their dongles to the backend via a terminal. Once the backend receives a user’s encryption key derived from one of the user’s OTPs, it decrypts their dongle’s encounter entries and verifies their consistency. If the entries are consistent, the backend adds the encounter entries to the risk database and/or the epidemiology database. The global risk information is broadcast to the users’ dongles via network beacons. Dongles use this information to compute a risk score based on the number of matched ephemeral ids and other features of each encounter encoded in the beacon broadcast. If the risk exceeds a certain threshold, the dongle notifies the user via a LED so they can self-isolate and get tested. For the case that users carry their dongles openly visible in public spaces, the LED can be temporarily deactivated in order not to accidentally reveal a potentially incoming risk notification to third parties. Refer to *Supplementary Information* Sections 1.4 and 2 for details on data upload and risk dissemination respectively.

#### Risk score calculation

Whenever a user dongle receives risk information from the backend, it updates the owner’s risk score locally (within the dongle). The individual risk score is proportional to the period during which the individual and diagnosed individuals were near the same beacons, as measured by the number of ephemeral ids contained in the risk information matching those stored in the dongle. Each ephemeral id may be weighted differently according to beacon-dependent parameters, such as indoor/outdoor, air quality, ventilation, and ambient noise. These parameters were stored by the dongle when it received the beacon’s transmission. How these features are weighted depends on parameters provided by the backend as part of the risk information. The parameters can be determined by the backend using machine learning techniques [28] and reflect the latest scientific knowledge about the disease.

#### Security and Privacy

PanCast’s security properties are comparable to or improve upon that of SPECTS [46, 47]. Its user devices transmit no information in normal operation and its dongles have a smaller attack surface than smartphones. While PanCast indirectly provides information about the locations where encounters occurred via the known beacon locations, this information is revealed selectively and only when users explicitly choose to do so (e.g., after the owner is tested positive). When an individual is diagnosed, they explicitly consent to the transmission and can select the information they wish to transmit from their device to the health authority. Even the most privacy-conscious users who disclose nothing and never transmit anything from their dongles (i.e., only listen passively) receive risk notifications arising from space-time proximity to diagnosed users who choose to disclose information. Moreover, a user learns about potential risks in a visited location only when a diagnosed individual visited the location within a certain time window, but does not otherwise learn the location history of positively tested individuals. Finally, the risk broadcast provides strong differential privacy guarantees for the number of risk entries and the number of diagnosed individuals contained in a risk broadcast. This ensures that an adversary, even one in possession of offine information about some users, learns nothing about the health status or whereabouts of the remaining users through the system. We provide a detailed discussion of PanCast’s security and privacy characteristics in Section 3 of the *Supplementary Information*.

### Epidemiological Simulations

#### Epidemiological model

To study and contrast the effects of PanCast in the context of generic SPECTS, we simulate various contact tracing policies using a spatiotemporal epidemic model recently introduced by Lorch et al. [28]. This agent-based model leverages POIs from OpenStreetMap [29] and population density data from Facebook [48] as well as information about household structure and age demographics to build a mobility model of a given region—here of Tübingen, Germany. Following the gravity model [49] as well as additional assumptions about the individual mobility behavior, the model simulates the mobility traces of individuals in the region. In particular, the model assumes that the probability of an individual visiting a specific site decreases with the distance between their household and the site. Refer to [28] for a detailed overview of the relevant parameters.

By modeling transmissions of the disease explicitly at the sites they occur, the model is directly applicable to simulating the effects of our proposed system and comparing it with existing SPECTS. The disease dynamics follow an extended SEIR compartmental model [50] and are similar to other SEIR variants used in the context of COVID-19 [43, 51]. Specifically, each individual is defined to be in one of several epidemiological states (e.g., susceptible, symptomatic, or resistant) at any given point in time, and individuals transition through these states by interaction with others at sites and in their households. Section 5.1 and Figure 1 in the *Supplementary Information* provide further details on the baseline disease dynamics of the model. Its underlying temporal point process framework explicitly represents events when individuals check in at POIs, get in contact with and infect each other, change epidemiological state, and are affected by or interact with TTI measures [52]. The model further captures non-contemporaneous infections, allows for variability in transmission rate at different POI types, and faithfully incorporates contact tracing measures at POIs, such as manual contact tracing.

#### Model parameters

Site and household transmission rates are estimated using Bayesian optimization as proposed by Lorch et al. [28]. Following previous studies, we set the proportion of asymptomatic individuals among all infected individuals in the population to 0.4 [43, 53, 54] and the relative asymptomatic transmission rate to 0.55 [51]. After an infectious individual leaves a site, we assume that their relative non-contemporaneous transmission rate to others decays with a half-life ten times shorter than estimated for aerosols under laboratory conditions [41]. We truncate the possibility of non-contemporaneous transmission when the relative transmission rate drops below 10%, which occurs 21 minutes after an individual leaves a site. The remaining disease parameters related to SEIR progression from having been exposed to recovery are taken from the literature [55–60] and follow the values stated in [28].

#### Contact tracing

Our simulations include manual contact tracing as well as digital contact tracing using either PanCast or SPECTS. Independent of the tracing method, whenever a contact of a diagnosed individual gets successfully traced, the infection risk is estimated by taking into account the duration of the contact and possibly environmental factors (only for PanCast). Individuals with infection risk above a certain threshold are quarantined for two weeks, get tested within the next 24 hours, and receive the outcome of the test 48 hours later. We choose the infection risk threshold to correspond to a 15 minute contact with a symptomatic individual in the model, which is in accordance with SPECTS currently employed in Germany, Switzerland, the United Kingdom, France, and Australia [61–65].

All simulations using PanCast or SPECTS also implement manual tracing by assuming that a proportion *p*_reachable_ = 0.5 of visitors leave their contact details at social, office and education sites, so that they can be reliably contacted, e.g., via phone. Upon receiving a positive test result, we assume that every individual participates in a manual contact tracing interview independent of their participation in digital tracing. In the tracing interview, we assume a person only remembers a fraction *p*_recall_ = 0.1 of their visit history of the past 14 days. This accounts for the fact that most people cannot remember every place they have visited and some people might not be willing to share any data or cannot be reached for an interview. This choice of the parameter values is conservative because PanCast directly benefits from larger values of *p*_recall_ and *p*_reachable_ via interoperation with manual tracing as opposed to SPECTS. If an infected person *i* recalls a visit at which they encountered a person *j* and individual *j* is reachable, the contact between *i* and *j* is traced successfully and *j* gets quarantined and tested if the exposure risk exceeds the previously stated 15 minute threshold. Here, we assume that individuals remember the time and duration of their visits at a 15 minute granularity, the one used by PanCast.

For SPECTS, we assume that a certain proportion *p*_digital_ of the population adopts the digital tracing technology. When an individual using SPECTS is tested positive, all contacts at POIs of the past 14 days that also use the system get traced. SPECTS and manual tracing do not interoperate but complement each other. Any person may still participate in manual contact tracing, but no contact information can be shared between the two systems as SPECTS do not operate using space-time information.

For PanCast, we likewise assume that the same proportion *p*_digital_ of the population has adopted the system and is carrying a user device (dongle or smartphone). We place beacons at a proportion *p*_beacon_ of POIs. This can be done at random or strategically, by taking into account quantities related to the site-specific probability of infection, e.g., by ranking the sites according to their integrated visit time. Whenever a person carrying a user device gets tested positive, all contacts that also carry user devices at sites with beacons get traced. In addition, the information can be used to trigger manual tracing actions at all sites registered by the device of the positive-tested individual (i.e., PanCast supports manual tracing). Likewise, when a person that tested positive does not carry a PanCast user device but participates in a manual contact interview and recalls a visit to a site with a beacon, all individuals carrying dongles at this site can be traced (i.e., manual tracing supports PanCast).

In our simulations, both for manual and digital contact tracing, we assume that the traced contacts are notified and quarantined instantaneously. Moreover, for manual contact tracing, we assume that a contact tracing interview takes place instantaneously upon receiving a positive test result. In practice, these assumptions may be violated and influence the effectiveness of both manual and digital contact tracing [4]. Section 5.2.1 and Figure 3 of the *Supplementary Information* show that a 24-hour delay of the manual tracing notifications does not qualitatively change our findings.

## Data Availability

Code and data to reproduce our epidemiological simulations are available at https://github.com/covid19-model/simulator/tree/beacon.

http://pancast.mpi-sws.org

## Data Availability

Code and data to reproduce our epidemiological simulations are available at https://github.com/covid19model/simulator/tree/beacon.

## Acknowledgements

We would like thank Nuria Oliver, Viola Priesemann, Nasim Rahaman, Peter Schwabe, Clara Scheidewind, Michael Meyer-Hermann, and the CIFAR contact tracing working group for helpful feedback.

## Author Contributions

G.B., P.D., D.G., M.G.R., A.M. and B.S. designed research; G.B., R.D.V., P.D., D.G., M.G.R., P.I., H.K., M.L., L.L, A.M. and B.S. performed research; H.K. and L.L. analyzed data; and G.B., R.D.V., P.D., D.G., M.G.R., P.I., H.K., M.L., L.L, A.M. and B.S. wrote the paper.

## Competing Interests

We declare no competing interests.

## Notes

### Competing Interest Statement

The authors have declared no competing interest.

### Funding Statement

No external funding was received.

### Author Declarations

All the data used in the study is publicly available and is anonymized.

### Summary of Updates

Additional results have been added, including Reff analyses, interoperability with existing systems, sensitivity analysis, stratified sensitivity-specificity. A review of existing smartphone-based contact tracing applications have been added. As a result, the Results section and Figures 2-4 have been significantly updated in the main and Sections 4-5 have been added to the Appendix.

