## Supplementary Information for "Listening to Bluetooth Beacons for Epidemic Risk Mitigation"

### Supplementary Information
### Listening to Bluetooth Beacons for Epidemic Risk Mitigation

Gilles Barthe[1] Roberta De Viti[2] Peter Druschel[2] Deepak Garg[2]
Manuel Gomez-Rodriguez[2] Pierfrancesco Ingo[2] Heiner Kremer[3]
Matthew Lentz[4] Lars Lorch[5] Aastha Mehta[2] Bernhard Schölkopf[3]

**1 Data Collection and Processing** **2**

**6 References** **23**

[1]Max Planck Institute for Security and Privacy, Bochum, Germany. [2]Max Planck Institute for Software Systems, Saarbrücken and Kaiserslautern, Germany. [3]Max Planck Institute for Intelligent Systems, Tübingen, Germany. [4]VMware Research, Palo Alto, USA. [5]ETH Zürich, Zürich, Switzerland. Correspondence should be addressed to (Manuel Gomez-Rodriguez) and (Bernhard Schölkopf).

### 1 Data Collection and Processing

#### 1.1 Beacon and dongle registration

When a dongle is registered, it receives a dongle id $d$, the backend's public key, an initial clock $C_d$ synced to real time, a secret key $sk_d$ and a list of one-time passwords (OTPs) from the backend. When a beacon is registered, it receives a beacon id $b$, the backend's public key, a location id $loc_b$ corresponding to the location where the beacon is supposed to be installed, an initial clock $C_b$ synced to real time, and a secret key $sk_b$ from the backend. The backend stores the registration data of the beacons and dongles in a device database. Specifically, the database contains entries of the form:

$$\{\text{device type}, \text{device id}\} \text{ :- } \{\text{secret key}, \text{initial clock}, \text{clock offset}, \text{location id}\}$$

where the device type is either beacon or dongle, and the location id exists only if the device type is beacon. The clock offset in the backend's database denotes any divergence between the local timer of a device and real time that is known to the backend.

Each dongle and beacon has a coarse-grained timer of 1-minute resolution ($t_d$ and $t_b$ respectively), which is set to the initial clock value provided by the backend, and subsequently increments once every minute. A device stores its timer value to local flash storage at intervals of fixed length $L$, called epochs. A non-persistent variable tracks the epoch id or the number of epochs elapsed since the device's start ($i_d$ and $i_b$ for the dongle and beacon, respectively). In practice, we suggest an epoch length of $L = 15$ minutes.

The secret key of a device is known only to the device and the backend. This key is used to mutually authenticate the device and the backend to each other, and to establish a secure channel between the two, whenever the two communicate with each other. The OTPs provided to a dongle are also given to the dongle's owner in human-readable form (e.g., printed on paper). These OTPs are used to mutually authenticate the owner and the dongle whenever the owner interacts with the dongle (e.g., to initiate data upload to the backend).

#### 1.2 Encounter data collection

A beacon generates a new ephemeral id every epoch. In the $i^{th}$ epoch $i_b$, the beacon $b$ generates an ephemeral id $eph_{b,i}$ that is computed as follows.

$$eph_{b,i} = hash(sk_b, loc_b, i_b)$$

Here $i_b = \lfloor (t_b - C_b)/L \rfloor$ and $hash$ is a one-way hash function. The beacon broadcasts its information as a tuple of the form $\{eph_{b,i},\ b,\ loc_b,\ t_b\}$, which is captured by nearby user dongles. Suppose a dongle $d$ encounters a beacon $b$ when the dongle's local timer is $t_d$, and the beacon's timer, epoch and ephemeral id are $t_b$, $i_b$, and $eph_{b,i}$, respectively. Then the dongle logs an entry in its persistent storage as a tuple defined as:

$$\{eph_{b,i},\ b,\ loc_b,\ t_b,\ t_d\}$$

Beacons may broadcast the ephemeral ids several times within an epoch. Dongles persist only one entry for each unique ephemeral id they receive.

#### 1.3 Data structure size and storage requirements

Dongles store encounter data as tuples of the form

$$\{eph_{b,i},\ b,\ loc_b,\ t_b,\ t_d\},$$

where $eph_{b,i}$ and $loc_b$ denote the $i$-th ephemeral id and location of beacon $b$ respectively, and $t_b$ and $t_d$ denote the local clocks of beacon $b$ and dongle $d$ respectively. Location identifiers $loc_b$ are 8 bytes in size. These bytes may be used to represent GPS coordinates or a hierarchical location naming scheme (e.g., `country.city.zip`, or `country.city.bus_id`). Beacon timer and epoch counters are 4 bytes in size and have a 1-minute resolution. Thus they can present time for 16 years, which is well beyond the expected lifetime of the beacons. Beacon identifiers $b$ are 4 bytes in size, and represent a simple global counter for all beacons. Thus, we can support roughly 4.3 billion beacons in the world. The hash in the ephemeral id is generated by computing SHA-256 of the inputs and taking the least significant 15 bytes of the result. Overall, each beacon broadcast ($\{eph_{b,i},\ b,\ loc_b,\ t_b\}$) is $15 + 4 + 8 + 4 = 31$ bytes in length.

Similar to beacons, dongle timers and epoch counters are 4 bytes each and have a 1-minute resolution, and dongle identifiers are 4 bytes in size. Thus, for each unique ephemeral id received from a beacon, a dongle stores 35 bytes of data. We store an encounter with a beacon if the dongle observes the beacon's ephemeral id broadcast for at least $E_{min}$ minutes, which we assume to be 5 minutes.

We assume that users encounter on average no more than one unique beacon ephemeral id every 5 minutes throughout the day (24 hours). Accordingly, dongles need to store data for 288 encounters in one day or 4032 encounters in a 14-day window (the infectious period for the current COVID-19 as determined by health experts), which requires roughly 138KB of persistent storage in a dongle. In reality, a dongle is very unlikely to encounter a distinct beacon ephemeral id every 5 minutes continuously for 14 days, so this estimate is conservative.

#### 1.4 Encounter data upload

Diagnosed individuals may share their encounter data with health authorities to enable dissemination of risk information to people who might have been in their vicinity. We first discuss the requirements for enabling data upload from users' dongles. Then, we describe different upload mechanisms that can enable users with different levels of technological access to upload their data.

##### 1.4.1 Requirements

(i) User dongles only have Bluetooth connectivity; therefore, they need to use a networked BLE node to upload their data to the backend.

(ii) Since dongles have a minimal user interface, users fundamentally need access to a separate device through which they can authorize their dongle to upload data in case of a positive test. This device, subsequently called a *terminal*, can be any device that has a graphical user interface, supports BLE, and has an Internet connection. A user needs to trust the terminal to send correct commands to their dongle. In case they wish to perform an upload of selected data (described below), they must also trust the terminal with the data stored on their dongle. The terminal can be part of a kiosk installed in a test center, clinic, or doctor's office, or a personal device (e.g., a smartphone or a computer) owned by the user or a care provider.

(iii) Users may be allowed to share their data *selectively*. For this, a terminal can be used to allow users to review the history stored in their dongles and select what subset of the information they wish to release to the backend.

##### 1.4.2 Upload mechanisms

Users typically visit a test center or a clinic for testing and receive their results offline via email or phone. If the test is positive, they may wish to upload their data. Users may also voluntarily contribute their data for epidemic analytics in the absence of a test. We present different mechanisms for data upload, which vary in the time when the data is uploaded and when it is actually released to the backend.

In all cases, patients identify themselves at the time a test is taken using the normal procedures in place for this purpose. Normally, their contact details are recorded along with the id of the test kit used for the patient. Once the test results are available, the user is informed using their contact details. If the result is positive, the notification includes a certificate that the patient has tested positive on the given date, which the user can forward to the PanCast backend.

**Delayed release** Here, a user initiates the data upload after they receive a positive test result. If the patient owns a smartphone or home computer, they can use it as a terminal for the upload. Users who do not own or have access to such a device and cannot leave their home to visit a terminal may use the smartphone of a person who visits them to provide essential services (e.g., delivering groceries).

When a user wishes to contribute their data, either because they have tested positive and wish to warn other people they have encountered or because they wish to contribute their data for epidemic analytics, they establish a secure connection to their dongle via the terminal. For this, the user enters one of the OTPs it shares with the dongle into the terminal. The dongle and the terminal, acting on behalf of the user, use this OTP as a common shared secret to authenticate each other and establish a secure connection. Once the connection is established, the user may enable the upload and attach their test certificate if applicable. The dongle then encrypts the data it stores with a symmetric secret key it shares with the backend, and uploads to the backend via the terminal, along with any certificate.

**Early release** Alternatively, users may choose to upload their data into escrow at the time of their testing, pending a positive test result. Here, the user establishes a secure connection to their dongle using a terminal at the testing site, and consents to contributing their data if the test result turns out positive. The data is immediately uploaded to the backend in encrypted form, using an encryption key derived from one of the OTPs the dongle shares with the user. The user is asked to reveal this OTP to the testing site, which uploads it to the backend along with the certificate if and when the result is positive.

With the early release approach, users trust the testing center to release their escrowed data only in case of a positive test result. Alternatively, users themselves can upload the OTP to release their escrowed information upon receiving a positive test result, for instance, through an automated phone system.

##### 1.4.3 Verification of encounter ephermeral ids

When a user uploads their data to the backend, the backend checks the validity of the provided information by running a consistency check. The consistency check verifies that the value of each ephemeral beacon id $eph_{b,i}$ is consistent with the time at which the dongle received it. It accommodates known clock offsets between encountered beacons and the dongle. We explain this in more detail below. We describe the handling of other forms of inconsistencies, for instance, due to misconfigured beacons or relay attacks in SI Section 3.2. Suppose the encounter happened at real time $T$ and recall that an encounter entry is of the form $\{eph_{b,i},\ b,\ loc_b,\ t_b,\ t_d\}$. Taking timer offsets into account, we have the following equation:

$$t_d + \delta_d = T = t_b + \delta_b.$$

Here, $\delta_d$ and $\delta_b$ are the clock offsets of the dongle and the beacon known to the backend. The backend checks that $t_d + \delta_d = t_b + \delta_b$. Next, the backend calculates the beacon's epoch id $i_b = \lfloor (t_b + \delta_b - C_b)/L \rfloor$ at the time of encounter. The backend retrieves the beacon's secret key $sk_b$ using the beacon identifier $b$ from the encounter entry, and computes the expected ephemeral id $hash(sk_b, loc_b, i_b)$ for the epoch $i_b$. If this expected ephemeral id matches the ephemeral id $eph_{b,i}$ in the encounter entry, then the encounter entry is consistent and the backend adds it to its risk database, else the entry is ignored.

### 2 Risk dissemination

In this section, we describe a protocol for risk dissemination. Such a protocol must balance privacy and efficiency concerns. On the one hand, the protocol must protect the location history and identity of diagnosed patients. On the other hand, the protocol must ensure that relevant risk information reaches potentially affected users' dongles in a timely manner. We opt for a protocol where network beacons broadcast global risk information periodically, and user dongles passively listen for relevant risk information whenever they are in proximity of such a beacon. With this approach, dongles receive risk information while revealing no information about their identity or their history to the backend, the network beacons, or to other users.

The risk information consists of a list of ephemeral ids. The ephemeral id of a beacon $b$ for epoch $e$ is included in the list if and only if a diagnosed individual encountered $b$ in epoch $e$. To reduce the bandwidth overheads and broadcast delays, the list of risk entries is limited to the period of contagion of the disease (e.g., 14 days in the case of COVID-19). The list can also be compressed using cuckoo filters [1], at the cost of a small percentage of false positives. For example, to ensure an average false positive rate of 0.01% for each user in a 14-day period, we require a cuckoo filter with entries of size just 27 bits (as opposed to the 15-byte ephemeral ids), which reduces the size of a risk broadcast by $\sim$4.4x. The risk information may additionally contain values of parameters relevant for risk score calculation performed on user devices, e.g., the weights of various features of an encounter in the risk estimation. All risk information is signed by the backend to allow detection of any tampering by intermediate nodes, including network beacons.

In the following, we discuss the need to add noise in the risk dissemination protocol and the efficiency of the entire protocol.

#### 2.1 Noising the risk dissemination broadcast

We present two scenarios where the *number of entries* in the risk broadcast could potentially reveal an individual's identity, location history, or health status to an adversary in the locality of the individual. These leaks arise without the adversary having even encountered the individual at all. We then describe our solution to mitigate such leaks.

(i) **Whereabouts of diagnosed individuals.** Suppose Alice learns from the local news that there was only one case of infection in the past few days within some geographic region. Furthermore, Alice happens to know that Bob was diagnosed, that he carries a dongle, and that he agreed to upload his encounter history when he got diagnosed. Now, if Alice learns that the risk information for her region includes a non-zero number of ephemeral ids, she can infer that Bob left his home at some point while he was contagious. Dually, if Alice sees that no risk information is provided for her region, she can infer that Bob has not been near a beacon within the region and during the period in question. In short, the length of a risk notification broadcast (zero vs. non-zero) reveals to an adversary information about the whereabouts of a diagnosed individual. Note that this problem exists even if a cuckoo filter is used to encode risk information, because the size of a filter optimized for space reveals information about the number of elements it includes.

This information leak arises only if the location history of diagnosed users is always uploaded to the backend. In practice, users have a choice to not upload their history to the backend. If users exercise this choice frequently, then an adversary cannot learn much from the absence of risk entries. Nevertheless, we recognize that, with what we have described so far, such a leak is possible.

(ii) **Health status of an individual.** Suppose Alice lives in an area with very few people, say $n$ people, and Alice is able to track the movements of $n-1$ of these people through outside channels. Suppose the risk information Alice receives contains more ephemeral ids than can be accounted for by the movements of the $n-1$ people Alice is tracking. At this point, Alice knows that the $n$-th person (whom Alice is not tracking) must be sick as well. Even though such an attack requires a significant amount of offline information and may be difficult to use in practice, it does raise privacy concerns.

| $\epsilon$ | $\delta$ | **Offset(t)** | **99%-ile noise** |
|---|---|---|---|
| 0.5 | 0.001 | 46638 | 78196 |
| 0.2 | 0.001 | 94981 | 173938 |
| 0.1 | 0.001 | 160148 | 318262 |
| 0.05 | 0.001 | 263153 | 580176 |

| $\epsilon$ | $\delta$ | **Offset(t)** | **99%-ile noise** |
|---|---|---|---|
| 0.5 | 0.01 | 28181 | 59850 |
| 0.2 | 0.01 | 49361 | 129117 |
| 0.1 | 0.01 | 70588 | 231983 |
| 0.05 | 0.01 | 90283 | 420116 |

Table 1: 99th percentile noise required for various values of $\epsilon$ and $\delta$. Sensitivity ($A$) is 4032.

Note that both these leaks rely solely on the *number of ephermeral ids in a risk notification.* We propose to mitigate these leaks by adding noise to the risk notification broadcast in order to hide the actual number of ephermeral ids. For this, we add junk ids that do not correspond to any real beacon and, therefore, do not match the history of any user dongle. Since our threat model assumes that no adversary can monitor the ephemeral ids from a significant fraction of beacons, no adversary can distinguish these junk ids from legitimate ids. The number of junk ids can be chosen to satisfy differential privacy for all individual users, which we describe next.

**Differential privacy.** Our goal is to add a random number of additional junk ids to a risk broadcast to make the total number of entries differentially private for all individual users. The number of junk ids we add must obviously be non-negative. For this, we adapt a mechanism proposed in prior work [2]. Given a risk broadcast, we add $N$ junk ids to it, where

$$N = t + \lfloor \tilde{X} \rfloor.$$

Here, $t$ is a natural number, and $\tilde{X}$ is a random value sampled from a Laplacian distribution with mean 0 and parameter $\lambda$ *truncated* to the interval $[-t, \infty)$. Note that $N$ is always non-negative. The values of $t$ and $\lambda$ we use depend on how much privacy we want. Specifically, to get $(\epsilon, \delta)$-differential privacy, we pick the following $\lambda$ and $t$.

$$\lambda \;=\; A/\epsilon \qquad\qquad t \;=\; \lceil \lambda \cdot \ln\left((e^{(A/\lambda)} - 1 + \delta)/2\delta\right) \rceil$$

Here, $A$ is the sensitivity of the risk broadcast function; it equals the maximum number of risk entries that could be contributed by a *single* diagnosed individual. Refer to the PanCast white paper [3] for a proof that this mechanism is $(\epsilon, \delta)$-differentially private.

SI Section 1 shows the 99th percentiles of the number of noise entries required for different values of $\epsilon$ and $\delta$. Here, we conservatively assume that $A$ is 4032 (see SI Section 1.3). In practice, the number of junk ids must be tuned for the smallest granularity of a region for which statistical information is available from other public sources. For instance, in Germany, noise should be added to the statistics of cities and towns since statistics in Germany are reported at this granularity.[1]

#### 2.2 Bandwidth requirements and broadcast time

We now discuss the anticipated bandwidth and time requirements for a network beacon to broadcast risk information of a certain size. We estimate the time to broadcast this risk information using the latest BLE (Bluetooth Low Energy) protocol v5.2 [4].

**Bandwidth.** As discussed in SI Section 1.3, we conservatively assume that a diagnosed individual records about 288 unique beacon ephemeral ids in a day ($N_{day}$). Furthermore, we assume that individuals are diagnosed at the very end of the infection window $W$; in other words, they upload their encounter entries for the last $W$ days. Thus, the number of risk entries from each diagnosed individual is $N = N_{day} * W$. Given $I$ diagnosed individuals, each having generated $N$ risk entries of size $S$ bytes each, the total number of bytes required for broadcasting all risk entries is $B = (I * N * S)$ bytes.

[1] `https://tinyurl.com/yxv74nep`

| Country | # new cases/day | # risk entries/day | Bytes/day (MiB) | Delay (s) |
|---|---|---|---|---|
| Australia | 18 | 390,838 | 1.258 | 13.191 |
| Germany | 3346 | 13,809,334 | 44.447 | 466.065 |
| Italy | 3821 | 15,724,534 | 50.612 | 530.703 |
| France | 16036 | 64,975,414 | 209.133 | 2192.92 |
| Brazil | 26429 | 106,879,990 | 344.009 | 3607.2 |
| USA | 48639 | 196,430,710 | 632.242 | 6629.54 |
| India | 72019 | 290,698,870 | 935.658 | 9811.09 |

Table 2: Bandwidth and time requirements for broadcasting risk information (organized as cuckoo filters) in different countries. We include the cost of 318,262 noise entries, which is the 99%-ile for differential privacy with $\epsilon = 0.1$ and $\delta = 0.001$. Column 2 contains the 7-day moving average of daily new cases as of 11 October 2020.

Bluetooth Low Energy (BLE) v5.2 supports a new form of communication: isochronous channels [4]. Isochronous channels support both connection- and broadcast-oriented schemes. Here, we use only broadcast-oriented channels. A broadcast isochronous group (BIG) can contain up to 31 individual broadcast isochronous streams (BISs); each stream transmits data as bursts of packets in events that are synchronized to a fixed time interval. Streams can be sequential or interleaved. Devices can join a BIG and choose to listen to one or more BISs. As per the specification, BISs transmitted on a physical channel of 2 Mbps (LE 2M PHY) can nominally achieve a goodput of 1.66 Mbps. In practice, interference from WiFi and channel errors may reduce this goodput. Assuming that we can practically achieve a goodput of at least 0.8 Mbps, the time to broadcast $B$ bytes in a single BIS is at most $Delay = ((B * 8)/(0.8 * 10^6))$ seconds.

**Broadcast estimates.** SI Section 2 shows the bandwidth and latency requirements based on daily new COVID-19 cases reported in different countries [5]. $S$ is 15 bytes and $W$ is 14 days. Here, we assume that the risk entries of each region are coded as a cuckoo filter of size equal to the number of risk entries (column 3), and with entries of size 27 bits, which corresponds to a false positive rate of 0.01% for users in a 14-day period. Moreover, we include 318,262 noise entries, the 99%-ile of noise needed to provide differential privacy with $\epsilon = 0.1$ and $\delta = 0.001$.

In the above, note that the estimates in SI Section 2 are conservative in two ways: the maximum number of risk entries uploaded by a single diagnosed individual and the amount of differentially private noise added. The latter is also affected by the maximum number of risk entries possible from a single individual. In practice, most users are unlikely to encounter 288 unique ephermeral ids every day (as this corresponds to seeing a new ephemeral id every 5 minutes).

**Optimizing broadcasts.** Users may be interested in the risk information of only a few specific regions, such as their neighbourhood and place of work, or their daily commute routes. We propose several optimizations to allow faster dissemination of relevant information to users. First, we organize the risk entries by regions of reasonable size (e.g., countries or cities) and transmit the entries of each region on a separate BIS stream. Second, we reorder the streams in different network beacons based on anticipated priorities of users in the region. For example, beacons at airports may broadcast the risk information of source and destination cities before the information of other regions, whereas beacons within a city may broadcast risk information of the city first, followed by that of neighboring cities, the state, the country, and so on. Finally, multiple network beacons can be placed in a location to parallelize risk broadcasts of different regions. Beacons can be connected to a common power source and configured to use non-overlapping frequency bands for broadcast streams. Each beacon then broadcasts the information of a few regions using broadcast streams in its frequency bands. User dongles can tune to specific beacons to receive risk information of only specific regions.

**Disregarded alternative: Actively querying risk information.** We considered another alternative for risk dissemination, whereby users actively query the backend directly for risk information of specific regions. However, in this alternative, users' queries are inevitably revealed to the backend, which can then potentially infer their location history. Hence, we disregard this design in favor of the completely privacy-preserving passive broadcast model. Note that, in our current design, a user can receive risk notifications without ever transmitting anything from their dongle.

### 3 Security and privacy

In this section we discuss PanCast's privacy and security properties. SI Section 3.1 describes the threats mitigated by PanCast and the assumptions necessary for PanCast's guarantees. SI Section 3.2 analyzes PanCast's security, while SI Section 3.3 discusses potential privacy-violating information leaks.

#### 3.1 Threat model

In the following we assess potential security threads and the corresponding protective measures of each component of PanCast. We also state the respective trust assumptions.

**Beacons.** *Beacons are operated by untrusted parties, may fail, be accidentally misconfigured, or be corrupted by malicious parties. However, we assume that only a small fraction of beacons is affected at any time.*

BLE and network beacons are managed by untrusted third parties, but they are registered and approved by the backend, run trusted firmware, and only accept firmware updates signed by the backend. Network beacons relay risk information from the backend to user dongles. This information is signed by the backend so dongles can detect unauthorized changes by corrupt beacons. For timely dissemination of risk dissemination (liveness), PanCast requires that most beacons transmit risk information without change.

BLE beacons are prone to four types of issues, which may lead to inconsistencies in data collection and subsequently incorrect risk notification and epidemiological analysis. First, beacons may fail; for example, they may run out of battery or may be rebooted at any time. Clocks in beacons may go out of sync due to failures and reboots, or due to clock drift. Second, a beacon may be installed in a location different from the one it was registered for; such misconfigurations may result from human error or deliberate acts. Misconfigured beacons can cause the backend to mis-identify the physical location of potential transmissions, hotspots, or superspreading events. Third, attackers could relay and re-broadcast a legitimate beacon's transmissions at a different location, even in near-real time. Relay attacks can cause the system to temporarily infer transmission risks that do not actually exist (false positives). Fourth, an attacker could obtain illegitimate access to a beacon and hack it or break into it physically. A compromised beacon allows the attacker to steal its secret key and replicate the beacon's broadcast, thus creating false positives as with relay transmissions above. We assume that such failures, misconfigurations, and attacks are sporadic and uncoordinated. The backend can detect misbehaving and failed beacons, and initiate a manual repair. PanCast's backend can also detect and correct inconsistencies in the collected data to a large extent. While a beacon is unavailable for any reason, transmissions in its vicinity may be missed by PanCast (false negatives).

We do not address side-channel leaks of beacon secrets, for instance, through power or electromagnetic radiations.

**Dongles.** *Dongles may fail or be corrupted by malicious parties. However, we assume that only a small fraction of dongles is affected at any time.*

Dongles are registered and approved by the backend, run trusted firmware, and only accept firmware updates signed by the backend. What information users must provide to register and obtain a dongle is subject to local policy and legislation. Minimally, evidence should be required to prevent any individual from registering many dongles. Requiring additional information such as names and contact details enables health authorities to actively trace individuals at risk, but raises privacy concerns that may limit adoption.

Dongles may fail and be physically compromised. Dongle failures (e.g., reboots, permanent failures) may cause false negatives due to missed beacon broadcasts while the dongle is unavailable or the user obtains a new one. A physical compromise may allow the attacker to steal the owner's data from the dongle and make up plausible encounter histories, thereby potentially causing some false positives, some false negatives or adding misleading information about user behavior.

Side-channel leaks of secrets from dongles are out of scope. Also, PanCast does not address leaks through the sizes and timing of location history uploads from dongles to the backend. In principle, these can be mitigated by adding chaff traffic to dongle uploads, and having users upload chaff data at random times.

**Backend.** *PanCast's backend service and its controlling authority are trusted to not leak or misuse user registration information, secret keys shared with devices, the encounter histories that users upload, and the backend's private key. The backend is also trusted to transmit risk information correctly to beacons.*[2]

**User terminals.** *Terminals are trusted to a limited extent.*

There are two cases. First, users may use their personal computing devices (computers or smartphones) as terminals. These computing devices are implicitly trusted by users. Second, users who do not own personal computing devices may use public terminals, or use the smartphone of a care provider. These terminals are trusted to a limited extent: (i) when such a terminal is used to select information on a dongle prior to upload, the terminal must be trusted to not leak that information; (ii) when such a terminal is used to send commands to a user dongle, the terminal is trusted to send the correct commands. Terminals are not trusted in any other way.

**Eavesdroppers.** *Any BLE device may receive and store beacon broadcasts. However, we assume that no party has the ability to continuously receive and collect the BLE transmissions of a significant fraction of beacons within a large, contiguous geographic region.*

BLE eavesdroppers can receive the transmissions of beacons they are close to and conspiring users may aggregate their observations. However, we assume that no party has the ability to continuously receive and collect the transmissions of a significant fraction of beacons within a large, contiguous geographic region. Malicious users and eavesdroppers may attempt to combine information obtained through PanCast with any amount of auxiliary information about a subset of users obtained through offline channels to try and violate the privacy of the remaining users.

#### 3.2 Security analysis

We analyze security risks posed by misconfigured devices and adversarial principals in PanCast, which may lead to inconsistent encounters. Inconsistent encounters may arise in three ways: (i) the clocks of beacons or dongles are out of sync with real time; (ii) a beacon is misconfigured and placed at a location different from where it is registered; or, (iii) an illegitimate beacon re-transmits a legitimate beacon's transmissions at a different location.

In the above, note that inconsistencies in beacons and dongles are irrelevant as long as there are no infections being reported. Next, we discuss mechanisms to identify and mitigate inconsistencies in encounters reported to the backend.

**Clock inconsistencies.** Encounters become inconsistent when an ephemeral id is found to have been used for more than one epoch length in real time. This may happen when devices crash and reboot after a long time, leading to encounter timestamps that are out of sync with real clock time. The backend can detect and fix such an inconsistency if it receives at least two encounters of an inconsistent device with consistent devices, where one encounter occurred before and the other after the device's crash. That is, the backend can fix an inconsistent beacon if it receives at least two encounters from consistent dongles with the beacon, and similarly, it can fix an inconsistent dongle if it receives at least two encounters of the dongle with consistent beacons. We now show an example of how the backend fixes beacon and dongle inconsistencies.

Suppose, at real time $T$, a dongle and a beacon encountered each other with local timers at $t_d$ and $t_b$ respectively and clock offsets at $\delta_d$ and $\delta_d$, respectively. Suppose the beacon crashes and reboots at real time $T+\tau$ where $\tau > L$. Without loss of generality, assume that the same dongle encounters the beacon after the beacon has rebooted. The beacon's timer after reboot is $t_b + 1$, while the dongle's timer is $t_d + \tau$. Assume that $t_b$ and $t_b + 1$ lie in a single epoch interval, i.e., the beacon's epoch id corresponding to the two times is

[2]As explained in SI Section 1.4, secret keys shared with user dongles are only used to establish secure sessions in which dongles upload user histories. Consequently, leaking keys shared with dongles is no worse than leaking uploaded user histories directly.

the same. The dongle's encounter history includes two entries of encounters with the beacon as follows:

$$\{eph_{b,i},\ b,\ loc_b,\ t_b,\ t_d\}$$
$$\{eph_{b,i},\ b,\ loc_b,\ t_b+1,\ t_d+\tau\}$$

When the backend observes the two entries from the dongle with the same ephemeral id but with the dongle's local timers more than one epoch length apart, the backend knows that the inconsistency was introduced due to a beacon restart. In this case, the backend updates the beacon's clock offset $\delta_b$ and the real time from when the offset comes into effect, and appends the tuple into the beacon's entry in the database. Specifically, the backend appends $\{C'_b, \delta'_b\}$ to a list of $\{\text{clock}, \text{offset}\}$ values stored with the beacon, where $C'_b$ is the beacon's global clock in the backend at the time of encounter, and

$$\delta'_b = ((t_d + \tau) + \delta_d - (t_b + 1)).$$

A similar mechanism can be used to detect inconsistencies due to the crash of a user dongle. If there are multiple entries where the difference in the dongle timestamps does not match with the difference in beacon timestamps, the backend would update the dongle's clock offset appropriately. A beacon or dongle is restored to a consistent state when its clock offset equals the difference between its clock and local timer values.

**Beacon misconfiguration.** Inconsistencies also arise if a beacon transmits information inconsistent with its location. Such inconsistencies can arise if (i) a beacon was (accidentally or maliciously) installed in a location different from where it is registered, (ii) a spoofed beacon configured with the secret key of a legitimate beacon re-transmits the same ephemeral ids in a different location, or (iii) an adversary replays the ephemeral ids of a legitimate beacon in other locations [6]. All cases lead to the same inconsistencies due to the fact that the spoofed beacon is in a location different from where it is expected. First, dongles that travel between a spoofed beacon and nearby, legitimate beacons will appear to have traveled implausibly long distances in a short period of time. The backend can detect this when such a dongle uploads its history. Second, users with GPS-enabled smartphones can directly observe the problem when they see a beacon transmission with a signed location that is different from the phone's current location by more than the BLE range. Such phones may report the inconsistency to the backend.

#### 3.3 Privacy analysis

Table 3 summarizes the information disclosed to different entities at various stages of PanCast's operation. Note that BLE beacons only transmit information unidirectionally and learn nothing about nearby users. In the following, we elaborate on the possible attacks by the remaining principals based on the information disclosed to them.

##### 3.3.1 Leaks to the backend

The backend learns the encounter histories of users who volunteer to upload this information by design. How much personally identifiable information (PII) the backend learns about users depends on the dongle registration policies of a given jurisdiction. From a technical standpoint, it suffices if the backend knows a pseudonym for each registered user and the associated dongle. In practice, some additional information may be required for Sybil mitigation, such as an email address, phone number, or other id.

The backend learns that the owner of a dongle is sick when the dongle uploads its encounter history and the associated test certificate. In addition, the backend learns a subset of the owner's recent whereabouts approved by the owner, at the granularity of beacon visits and epochs. Additionally, the backend learns the whereabouts of volunteer users who have opted to share their encounter history even when not sick. The backend can also infer an over-approximation of the possible social contacts among sick users and volunteers at the granularity of beacon visits and epochs.

| Operation stage | Backend | Other users | Network beacon | Terminal |
|---|---|---|---|---|
| Registration | PII (e.g., phone #), user-dongle mapping, beacon-location map, devices' secret keys, | none | none | none |
| Encounter logging | none | none | none | none |
| Encounter upload | location history | none | none | upload size, location history* |
| Risk notification | none | common subset of location history, but anonymized** | noised broadcast size*** and time | none |

Table 3: Information about a user disclosed to other entities during PanCast's operation. Notes: *Only if user wishes to use the terminal to select data to upload. **Other users only learn a list of (location, time) pairs where they intersected with at least one sick user. There is no information about how many users were sick, or their identities. ***Beacons only observe differentially-private (noised) broadcast size.

##### 3.3.2 Leaks to other users

In PanCast, user dongles learn nothing about other users except through the risk notifications. Therefore, it is impossible for users to learn information about other healthy users through the system. Moreover, users can maximally learn information about diagnosed users that those users have volunteered to upload. As described in SI Section 2.1, PanCast provides differential privacy in the number of risk entries from diagnosed users and the number of diagnosed users itself.

However, we note that this measure is effective only against adversaries whose history does not match any of the risk information entries, i.e., against users who were not in the vicinity of a diagnosed patient who uploaded their history. If a diagnosed user Alice uploaded their data to the backend, a user Bob who was recently near a beacon at the same time as Alice may be able to identify Alice or learn about her whereabouts from the risk dissemination information. Here, we describe two scenarios in which a user can learn some information about a diagnosed individual based on the risk information broadcast.

**Location history of a diagnosed individual.** Suppose Alice learns from the local news that there was one case of infection in the past few days, and she receives the associated risk information. If some ephemeral ids in the risk notification match Alice's recorded ephemeral ids, Alice learns a partial location history of the infected individual. Furthermore, by intersecting the common locations visited by the patient with her out-of-band observations of who was present, Alice may be able to identify the individual or narrow the list of candidates.

*Such a leak is inherent in any risk notification system, including decentralized SPECTS, in which user devices compute their own risk score based on encounters with a patient.* However, only users who have visited some of the same beacons around the same time as a sick person can benefit from the leak. To exploit the leak to learn the identity and whereabouts of arbitrary sick people, an attacker would require the ability to collect the ephemeral ids transmitted by all beacons within a geographic region of interest, which we believe is largely impractical.

**Identity of a diagnosed individual.** Suppose Alice visited a location at a time when there was only one other individual Bob around. Thus, the ephemeral id of the nearby beacon is recorded only in the dongles of Alice and Bob. If the ephemeral id in question appears in a risk notification, and Alice did not report sick, then she knows that Bob reported sick. This leak arises when only two (or a small number) of people were present at a given time at a given place. Note that such a leak is also inherent to all digital contact tracing systems, centralized or decentralized.

###### 3.3.3 Leaks to network beacons

As long as dongles listen to broadcasts of network beacons passively (which is sufficient for PanCast's operation), network beacons cannot learn anything about the presence or identity of the dongle.

###### 3.3.4 Leaks to user terminals

When a dongle uploads data to the backend via a terminal, the terminal can observe the size of the upload. The terminal cannot observe the content of the (encrypted) communication nor does it learn the identity of the dongle or user. Since the size of an upload could reveal how many beacons a dongle has recently visited, uploads can be padded to obfuscate this information.

If the user uses the terminal to select records prior to upload, then the terminal learns the part of the user's history that is stored on the dongle at the time of selection. To mitigate this concern, a user can use a trusted terminal for this purpose, e.g., their own personal computer or smartphone if they have one.

###### 3.3.5 Leaks from dongles

User dongles transmit data only when uploading information to the backend via a terminal, or to a terminal in case the user wants to select records. In both cases, the data is transmitted from the dongle only over a secure connection, and only after the dongle authenticates the other endpoint. Otherwise, retrieving the information stored in a dongle requires physical access.

### 4 Smartphone-based contact tracing applications

To expedite contact tracing and assist health officials in responding to patients and at-risk individuals, many digital contact tracing systems have been deployed. In each system, individuals install a smartphone application, which uses different technologies, such as Bluetooth, GPS, or QR codes, to track the user's trajectory and/or proximity to other users. We briefly discuss the different types of applications and their adoption statistics.

#### 4.1 Bluetooth-based applications

The Bluetooth-based applications record instances of physical proximity with devices of other individuals via close-range Bluetooth exchanges, referred to as encounters [7–13]. (We collectively refer to these applications as smartphone-based, pairwise encounter-based contact tracing systems or SPECTs.) When an individual is diagnosed with the infectious disease, they report their recent encounter history to a health authority, who may then use the encounter history to notify other infected or at-risk individuals who might have come into contact with the diagnosed individual during the individual's period of contagiousness (indicated by mutual encounters).

Depending on how risk status is determined and delivered to users, current SPECTS are broadly grouped into two types: centralized and decentralized. In a centralized system, the central health authority computes the risk status for all users in the system, and either broadcasts it to everyone (e.g., PEPP-PT-NTK [7] framework, systems based on the BlueTrace protocol [14] like OpenTrace [8] or TraceTogether [9]) or returns the user-specific risk status to an individual upon inquiry (e.g., ROBERT [10]). In a decentralized system, the health authority broadcasts information about patients' encounters, and the smartphone app computes the user's risk status locally on the device and reports it to the user. Examples of decentralized frameworks include those based on DP3T [11], PACT [12], and the TCN coalition [13].

In centralized systems, all user data is collected and managed centrally, which places a high degree of trust in the central authority. On the other hand, decentralized systems aim to preserve the privacy of users and, therefore, minimize data that is shared with authorities. However, this data minimization also prevents aggregation of data for epidemiological analysis.

#### 4.2 QR-based applications

In addition to SPECTs, QR code based systems have also been designed for digital contact tracing [15–18]. Organizations register with a central authority and receive a QR code which they install at their physical establishments (e.g. restaurants). When a user visits the establishment, they scan the code with a QR code scanner (typically a smartphone app). In centralized systems [16, 17], the application uploads information about the user's visit to a backend. In decentralized systems [15], the application just stores information about the user's visit in the user device. QR code based centralized systems have been deployed in Singapore [16] and, to a small scale in, Canada [17]. The current QR code based systems use static QR codes in each location. A static QR code corresponds to a beacon with a static id, which does not change through epochs, instead of an ephemeral id. However, ultimately, the use of a static id entails weaker privacy guarantees. For instance, static ids allow linking a user's multiple visits to the same locations, thus revealing more information about a user's location history. Furthermore, as a static id's link to a specific location is valid at all times, a user might record an id while visiting a place, but report it to the application at a later time. Such delayed reporting might alter the matching of the risk notification entries, enabling a user to learn information they would not have had access to otherwise.

#### 4.3 Adoption levels across countries

Each of the above applications and frameworks has seen varying levels of adoption in different countries. In Table 4, we present some of the most widely downloaded SPECT apps in different countries along with an estimation of their adoption level based on the number of reported downloads. Except Singapore, New

| App Name | Country | Framework | Downloads | Population |
|---|---|---|---|---|
| TraceTogether [22] | Singapore | BlueTrace | 4.2M+ | 5.7M |
| Aarogya Setu [23] | India | – | 169M+ | 1300M+ |
| NZ COVID Tracer [24] | New Zealand | DP3T/GAEN + QR | 2M+ | 5M+ |
| COVIDSafe [25] | Australia | DP3T/GAEN | 6M+ | 25M+ |
| Corona-Warn-App [26] | Germany | DP3T/GAEN | 25M+ | 83M |
| Immuni [27] | Italy | DP3T/GAEN | 10M+ | 60M |
| TousAntiCovid [28] | France | ROBERT | 5M+ | 67M+ |
| COVID Alert [29] | Canada | DP3T/GAEN | 6.2M+ | 38M |
| Canatrace [17] | Canada (Ontario) | QR code | 0.6M | 14M+ |
| SafeEntry/Sinpass [16] | Singapore | QR code | 1M+ | 5.7M |
| ViruSafe (PathCheck) [30] | Bulgaria | GPS | 50K+ | 7M |
| SafePlaces (PathCheck) [31] | US | GPS + BLE | 10K+ | 330M |

Table 4: Adoption of SPECTS in different countries

Zealand and Australia, the adoption of these apps in most countries is less than 25%. Note that the estimates are conservative as they indicate only the number of downloads and not actual usage. These estimates indicate that the existing SPECTS or other apps have not seen widespread adoption and, consequently, not proven as effective as one would have wished for [19–21].

### 5 Epidemiological Simulations

#### 5.1 Disease model dynamics

In this section, we provide a supplementary overview of the disease dynamics underlying the epidemiological model that we use to perform our simulations [32]. As already indicated in *Materials and Methods*, the model simulates mobility traces for each individual in a population $\mathcal{V}$ and across a set of sites $\mathcal{S}$. This is done by leveraging real data about POIs, population density data, household structure and age demographics. In turn, these mobility traces define an indicator function $P_{i,k}(t)$ that equals 1 if and only if individual $i \in \mathcal{V}$ is present at site $k \in \mathcal{S}$ at time $t$, and 0 otherwise. TTI affect the epidemiological model by amending the mobility traces of individuals—for example, by temporarily setting $P_{i,k}(t) = 0$ after tracing the contact with a positively tested individual.

Given the mobility traces, the disease dynamics follow an extended SEIR compartmental model [33]. By interaction with other individuals at sites and in their households, each individual may transition through a sequence of possible epidemiological states. Specifically, each individual is defined to be in one and only one of the following states at any given time: susceptible, exposed, asymptomatic, pre-symptomatic, symptomatic, hospitalized, resistant, or dead. Figure 1 illustrates the possible state transitions for a single individual in $\mathcal{V}$.

The instantaneous rate of exposure events for a given individual—directly related to the probability of exposure of an individual—is modeled by a time-variant intensity function that depends on the contacts with other individuals by way of the mobility traces. More specifically, Lorch et al. [32] propose to model the rate of exposure of individual $i \in \mathcal{V}$ across all sites $\mathcal{S}$ as

$$\lambda_i^*(t) = \sum_{k \in \mathcal{S}} \beta P_{i,k}(t) \sum_{j \in \mathcal{V} \setminus \{i\}} \int_{t-\delta}^{t} K_{j,k}(\tau) P_{j,k}(\tau) \; \gamma e^{-\gamma(t-\tau)} \, d\tau \tag{1}$$

where $K_{j,k}(\tau) = 1$ if individual $j$ is symptomatic or pre-symptomatic, $K_{j,k}(\tau) = \mu < 1$ if $j$ is asymptomatic, and 0 otherwise at time $\tau$. Following previous studies, we set the proportion of asymptomatic individuals among all infected individuals in the population to 0.4 [34–36] and the relative asymptomatic transmission rate to $\mu = 0.55$ [37]. The exposure rate in Eq. (1) captures non-contemporaneous infections in a time window of length $\delta$ and models an exponential decay of infectiousness after an infectious individual leaves a site. We set $\delta$ to roughly 21 minutes, which truncates the possibility of non-contemporaneous transmission when the relative transmission rate drops below 10%. As explained in *Materials and Methods*, we specify $\gamma$ by assuming that the relative non-contemporaneous transmission rate to others decays with a half-life that is ten times shorter than estimated for aerosols under laboratory conditions [38].

The remaining state transitions follow log-normal distributions as stated in [32] and are taken from the existing COVID-19 literature [39–44]. The base transmission rate at sites $\beta$ and analogously in households are estimated using Bayesian optimization as proposed by Lorch et al. [32]. More specifically, given the simulated mobility traces, the transmission rates are estimated by matching the simulated number of positively tested individuals with real COVID-19 case data over a period of eight weeks early in the pandemic.

#### 5.2 Additional results

In the following, we provide additional experiments and analyses of the experiments presented in the main paper. In Section 5.2.1, we provide a sensitivity analysis of the parameters of manual contact tracing in addition to complementing experiments. Section 5.2.2 provides additional experiments for the scenario of site-dependent transmission rates and finally Section 5.2.3 concludes with additional experiments on the interoperability of bluetooth-beacon-based systems like PanCast with SPECTS.

##### 5.2.1 Interoperation with manual tracing can improve efficacy at low adoption levels

Throughout this subsection we consider the scenario in which transmission rates are independent of site types, therefore PanCast and SPECTS have access to the same amount of information, thus eliminating one of PanCast's key benefits.

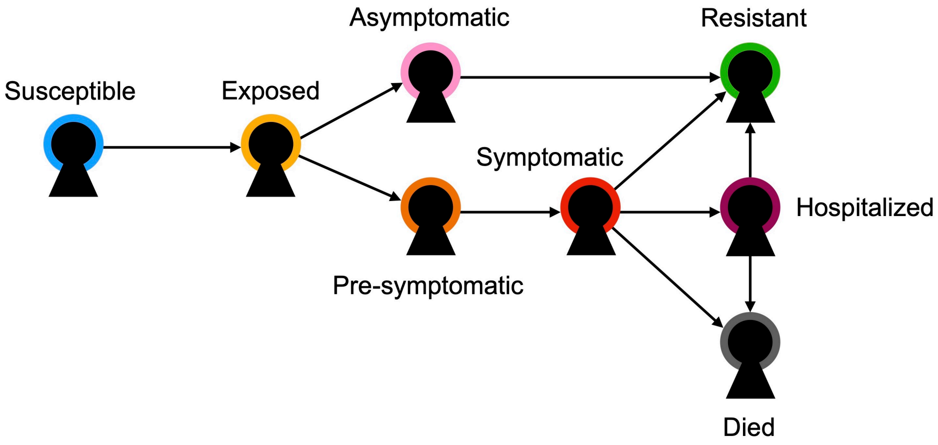

Figure 1: Epidemiological states and transitions of each individual in the disease model of the simulations [32]. The event-based state transitions are modeled using temporal point processes. In particular, for each individual in the model, the transition from *Susceptible* to *Exposed* is captured using a time-variant transmission rate that depends on the contact with infectious individuals. TTI influence the mobility patterns and hence the contacts of individuals, thereby affecting the probability of exposure.

**Manual contact tracing parameter sensitivity analysis** As discussed in the simulator section of *Materials and Methods* our implementation of manual contact tracing involves two parameters: $p_{\text{recall}}$ which quantifies the probability with which a positively tested individual recalls each of their visits to sites during a manual tracing interview and $p_{\text{reachable}}$ which describes the probability with which contacts can be notified via manual channels, i.e., phone or email. To keep the number of free parameters at a minimum, we assume that every positively tested individual participates in a manual contact tracing interview. The possibility of individuals not being compliant can be absorbed into a lower value for $p_{\text{recall}}$. We further assume that individuals can only be reached via manual channels if the contact with an infected person happens at a social, education or workplace site, as authorities typically only collect contact information at such sites, not at bus stops or grocery stores. Figure 2 investigates the sensitivity of the tracing probability with respect to these parameters, where the tracing probability is defined as the probability with which a random contact can be traced via manual tracing in combination with either PanCast or SPECTS. Figure 2a) shows the tracing probability as a function of the adoption of the digital tracing system and the manual tracing probability, i.e., the product of $p_{\text{recall}}$ and $p_{\text{reachable}}$. We observe that for intermediate values of $p_{\text{adoption}}$ and $p_{\text{recall}} \cdot p_{\text{reachable}}$ PanCast with 100% beacons shows an increased tracing probability compared to SPECTS. With decreasing proportion of beacons, PanCast looses its advantage and becomes less effective. Figures 2b)–2d) show the tracing probability in dependence on $p_{\text{recall}}$ and $p_{\text{reachable}}$ for different adoption levels of the digital system. The results suggest that by interoperation with manual contact tracing PanCast is capable of achieving tracing probabilities of a similar level as SPECTS even at beacon proportions of 25% or below. Our choice of $p_{\text{recall}} = 0.1$ and $p_{\text{reachable}} = 0.5$, which we use in all experiments with manual contact tracing, is located in the low tracing probability regime in the bottom left corner.

**Effect of delayed manual contact tracing** In the rest of the paper, we assume that manual contact tracing is happening instantaneously—upon receiving a positive test result all contacts of an individual that can be traced are immediately notified about their infection risk. While this assumption is justified for digital contact tracing, manual contact tracing requires an institution to notify potentially infected individuals manually via phone or email and hence is necessarily subject to delays. We investigate the effect of such delays by assuming that whenever a person has to be reached manually, we impose a delay of 24 hours before actions are being taken. This is the case either when a person is traced completely manually or via PanCast, when the infector is a user of the system but the contact is not and therefore has to be

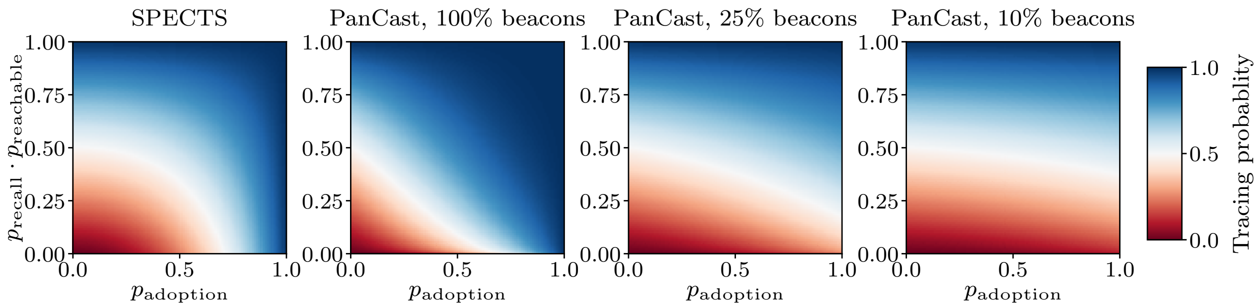

(a) Tracing probability as function of adoption of the digital systems and probability that a contact can be traced manually

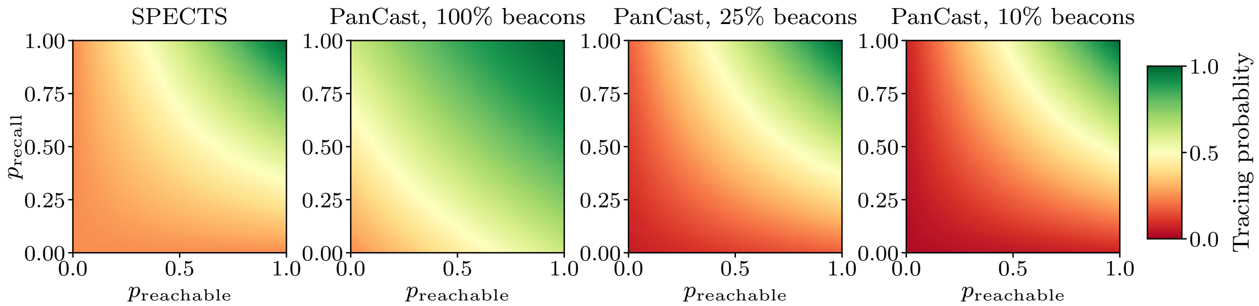

(b) 50% adoption of digital tracing

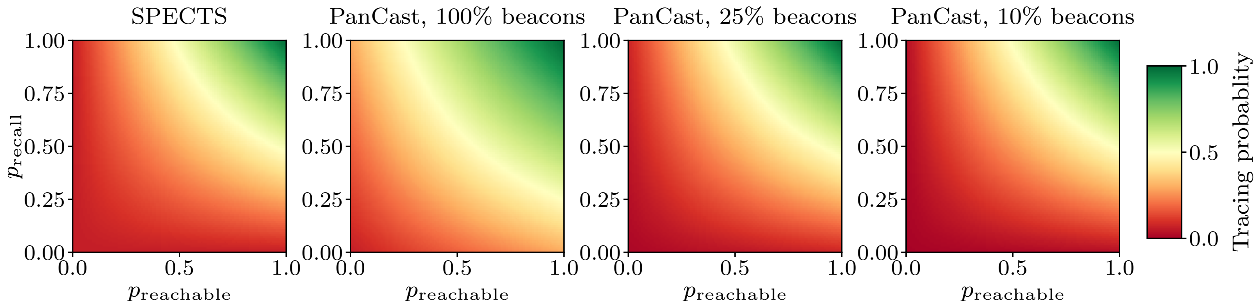

(c) 25% adoption of digital tracing

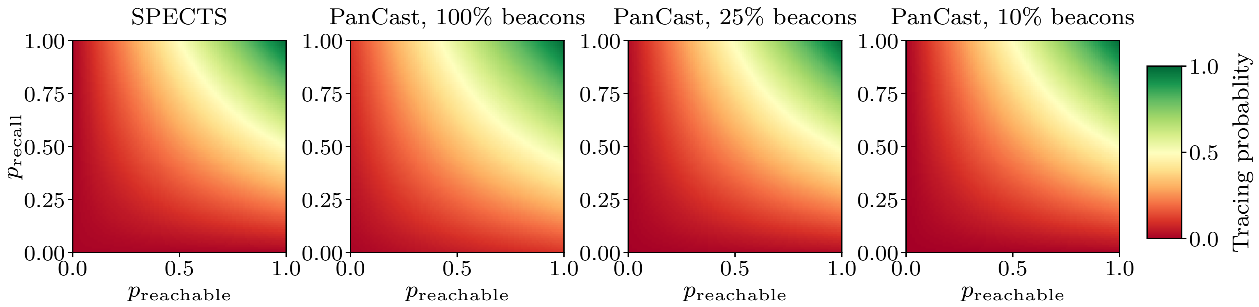

(d) 10% adoption of digital tracing

Figure 2: Dependency of tracing probability on manual contact tracing parameters. The tracing probability is defined as the probability with which a random contact can be traced via manual contact tracing in combination with either SPECTS or PanCast. Figure a) shows the tracing probability as a function of the adoption level of the digital tracing technology $p_{\text{adoption}}$ and the probability that a contact can be traced manually $p_{\text{recall}} \cdot p_{\text{reachable}}$. Figures b)–d) show the tracing probability as a function of the manual tracing parameters for different adoption levels of the digital systems.

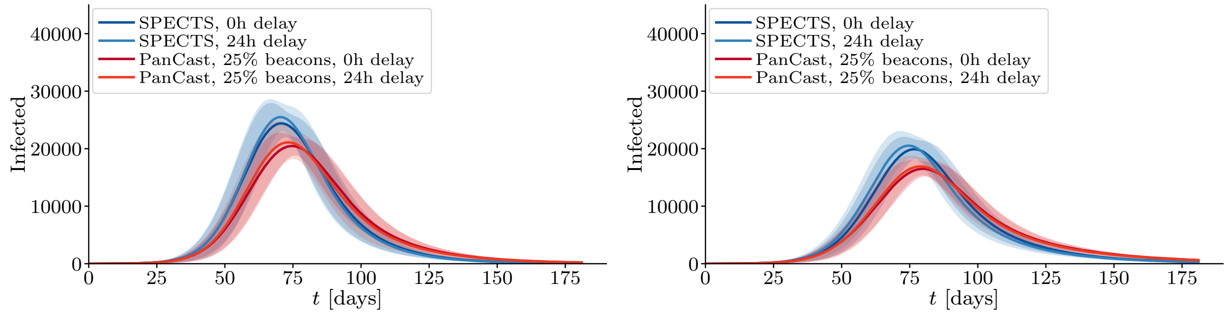

(a) 25% adoption

(b) 50% adoption

Figure 3: Effect of delayed manual contact tracing. Whenever a person needs to be notified about their infection risk manually, we assume a delay for the notification. The plots show the effect of a 24 hours delay in presence of PanCast or SPECTS for two different levels of adoption of the digital systems.

notified manually. Figure 3 shows the results of using a 24 hours delay for 25% and 50% adoption of the digital system respectively. We observe that while the delay causes slightly higher numbers of infected in both scenarios, with PanCast and with SPECTS, the difference seems to be reasonably small to justify our assumption of instantaneous notifications.

**Digitial contact tracing without manual contact tracing and with site-independent transmission rates** Figure 4 shows the results of different experiments in which either PanCast or SPECTS is employed without manual contact tracing. Unsurprisingly, Figures 4a) and 4b) show that when PanCast beacons are not employed at all sites, SPECTS strictly outperforms PanCast across all adoption levels in terms of reduction of infections and effective reproduction number. This is to be expected because PanCast trades a potential lack of beacons at some sites with the possibility to interact with manual tracing and utilize environmental information. Deprived of both of these advantages, PanCast shows worse performance, while converging to the performance of SPECTS in the case of beacons at all sites. The effective reproduction number, shown in Figure 4c) is consistent with this observation.

###### 5.2.2 Utilization of environmental information improves tracing decisions

For the experiments shown in Figures 3a) and 3b) in the main paper, we have set the global isolation budget, i.e., the number of individuals that can be quarantined simultaneously at any time purely on the basis of contact tracing decisions, to 10% of the population. The results shown in Figure 5 explore the effect of using different isolation budgets. We observe that PanCast with beacons at 25% of the sites keeps its advantage over SPECTS even for lower isolation budgets. However, we see that both tracing strategies become significantly less effective with smaller budgets. In particular, with a budget of 2% of the population, PanCast requires adoption levels above 25% and SPECTS above 50% to show a significant impact.

###### 5.2.3 Interoperation between SPECTS and beacon-based systems

Since the outbreak of the COVID-19 pandemic, a large range of contact tracing technologies have been developed and employed in several countries (see Section 4 for a short overview). While these systems can be regarded as separate entities, interoperation between different systems could provide a great potential to increase the overall efficacy of contact tracing efforts. To this aim, we investigate the effects when a generic SPECTS integrates PanCast's beacons and protocols, thereby gaining access to information collected in manual contact tracing as well as environmental information. Here, we exemplarily focus on the former aspect of interoperation with manual contact tracing, assuming equal transmission rates across site types. A similar analysis may be conducted for the aspect of environmental information. Figure 6a) shows the

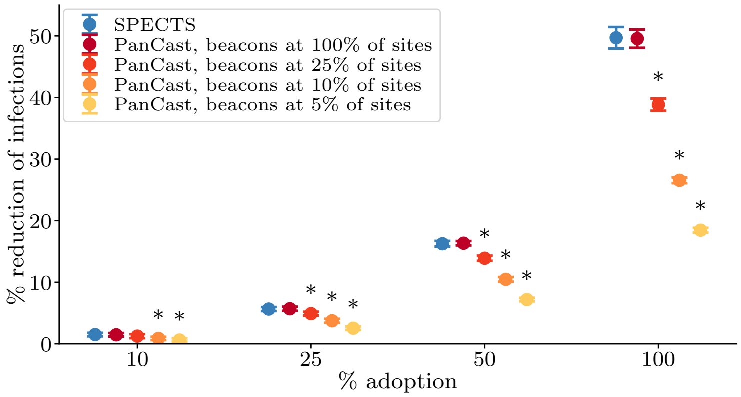

(a) Reduction of infections

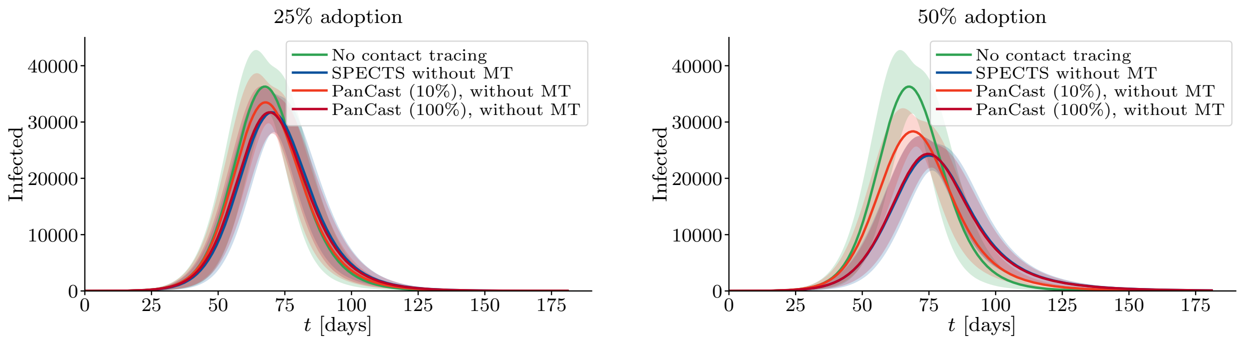

(b) Infected over time

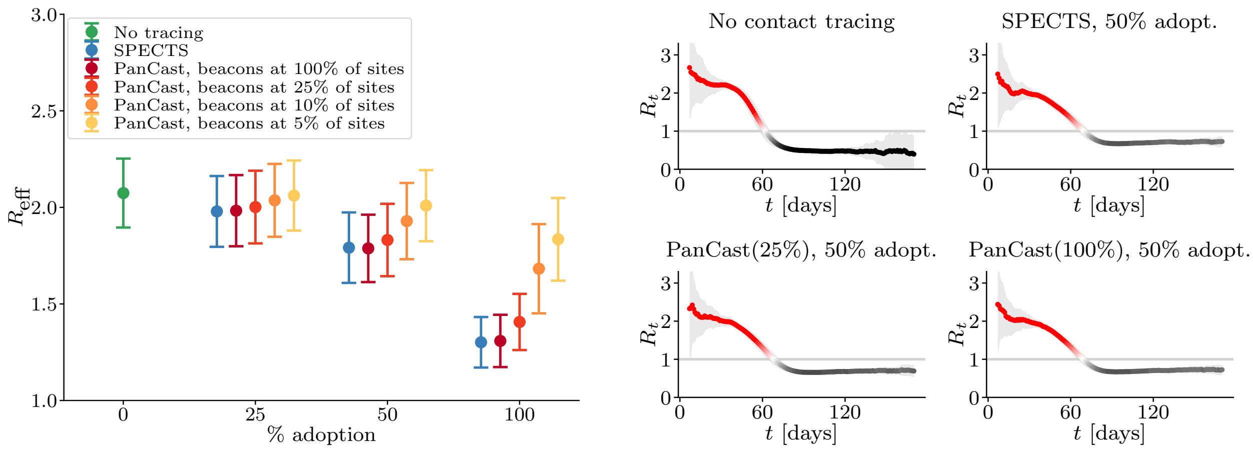

(c) Effective reproduction number

Figure 4: Efficacy of tracing systems without manual contact tracing and environmental information. Deprived of its advantages of incorporating manual contact tracing and environmental information, PanCast with incomplete beacon placement is outperformed by SPECTS while converging to the same efficacy for beacon proportions approaching 100%. Lines and points represent averages of 100 random roll-outs of the simulation, error bars correspond to plus and minus one standard deviation and, in figure a), the sign * indicates a statistically significant difference (two-sample t-test; p-value $< 0.05$) between PanCast and SPECTS.

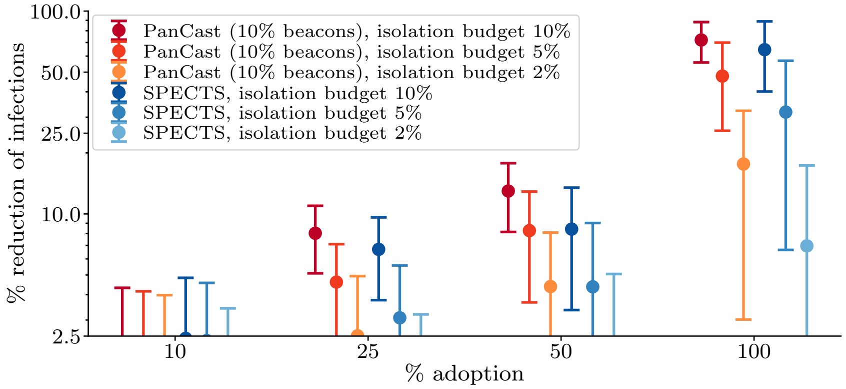

Figure 5: Utilizing site information under different isolation budgets without manual contact tracing. The budget is defined as the proportion of the population which can be quarantined purely on basis of tracing decisions. Positively tested individuals and their household members do not count towards this number.

reduction of infections achieved by SPECTS with integrated beacons. We observe that already for small proportions of beacons, SPECTS shows an increased utility, which further improves with increasing numbers of beacons. The results suggest that the same reduction of infections that is achieved by basic SPECTS at an adoption level of 50% can already be achieved at a lower adoption level of 25% by integrating beacons at 25% of sites. The left panel in Figure 6c) shows the effective reproduction number during the first weeks of the simulation before herd immunity becomes significant. Again, we observe that by better utilization of manual contact tracing information, SPECTS with integrated PanCast beacons appears to be capable of significantly reducing the effective reproduction number compared to a system that cannot leverage such information.

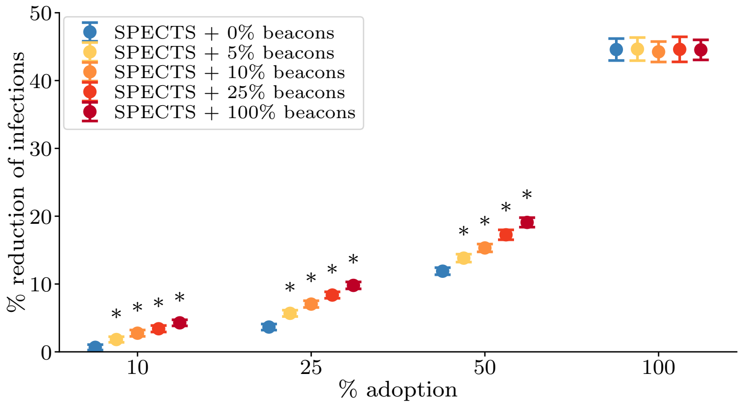

(a) Reduction of infections

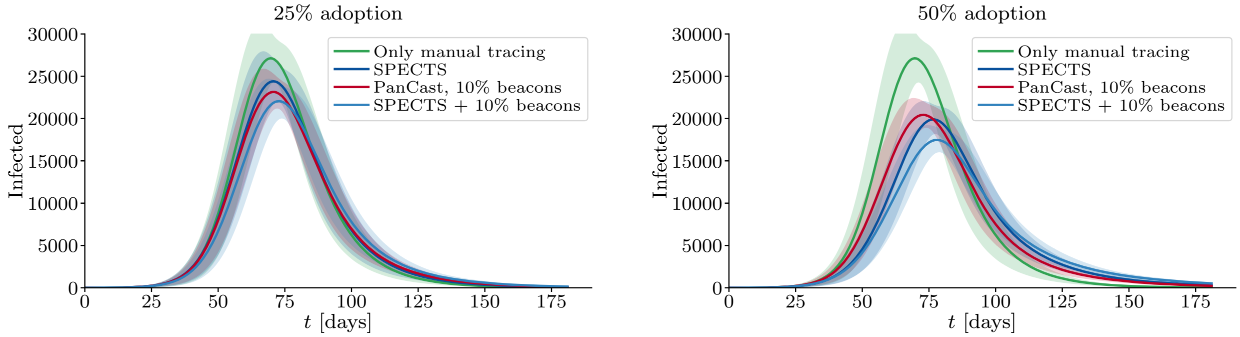

(b) Infected over time

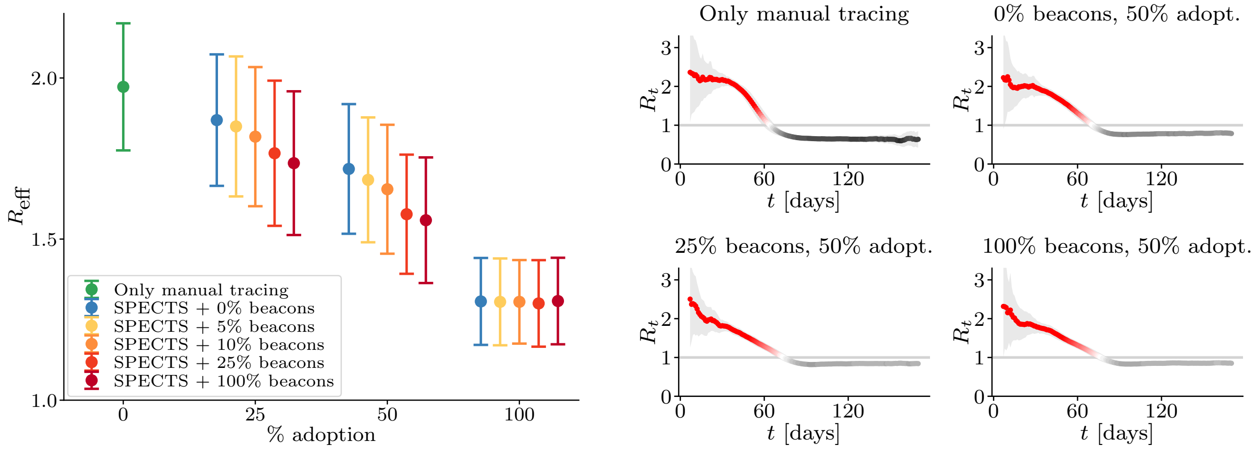

(c) Effective reproduction number

Figure 6: Interoperation between SPECTS and beacon-based systems help compensate low adoption. Figure a) shows the reduction of infections achieved by SPECTS that utilize manual tracing information provided by beacons. The sign * indicates statistically significant differences (two-sample t-test; p-value $< 0.05$) between PanCast and SPECTS. Figure b) shows the number of infected over time for different contact tracing strategies. The left panel in Figure c) shows the average effective reproduction number averaged over the first weeks of the simulation before the effects of herd immunity become significant, which we define as the time at which more than 10% of the population is not susceptible anymore. The right panel shows the reproduction number over time for several scenarios. Lines and points represent averages of 100 random roll-outs of the simulation, error bars correspond to plus and minus one standard deviation.
